# Genome-wide Association Study of Long COVID

**DOI:** 10.1101/2023.06.29.23292056

**Authors:** Vilma Lammi, Tomoko Nakanishi, Samuel E. Jones, Shea J. Andrews, Juha Karjalainen, Beatriz Cortés, Heath E. O’Brien, Brian E. Fulton-Howard, Hele H. Haapaniemi, Axel Schmidt, Ruth E. Mitchell, Abdou Mousas, Massimo Mangino, Alicia Huerta-Chagoya, Nasa Sinnott-Armstrong, Elizabeth T. Cirulli, Marc Vaudel, Alex S.F. Kwong, Amit K. Maiti, Minttu Marttila, Chiara Batini, Francesca Minnai, Anna R. Dearman, C.A. Robert Warmerdam, Celia B. Sequeros, Thomas W. Winkler, Daniel M. Jordan, Lindsay Guare, Ekaterina Vergasova, Eirini Marouli, Pasquale Striano, Ummu Afeera Zainulabid, Ashutosh Kumar, Hajar Fauzan Ahmad, Ryuya Edahiro, Shuhei Azekawa, Long COVID Host Genetics Initiative, FinnGen, DBDS Genomic Consortium, GEN-COVID Multicenter Study, Joseph J. Grzymski, Makoto Ishii, Yukinori Okada, Noam D. Beckmann, Meena Kumari, Ralf Wagner, Iris M. Heid, Catherine John, Patrick J. Short, Per Magnus, Karina Banasik, Frank Geller, Lude H. Franke, Alexander Rakitko, Emma L. Duncan, Alessandra Renieri, Konstantinos K. Tsilidis, Rafael de Cid, Ahmadreza Niavarani, Teresa Tusié-Luna, Shefali S. Verma, George Davey Smith, Nicholas J. Timpson, Mark J. Daly, Andrea Ganna, Eva C. Schulte, J. Brent Richards, Kerstin U. Ludwig, Michael Hultström, Hugo Zeberg, Hanna M. Ollila

## Abstract

Infections can lead to persistent or long-term symptoms and diseases such as shingles after varicella zoster, cancers after human papillomavirus, or rheumatic fever after streptococcal infections^1, 2^. Similarly, infection by SARS-CoV-2 can result in Long COVID, a condition characterized by symptoms of fatigue and pulmonary and cognitive dysfunction^3–5^. The biological mechanisms that contribute to the development of Long COVID remain to be clarified. We leveraged the COVID-19 Host Genetics Initiative^6, 7^ to perform a genome-wide association study for Long COVID including up to 6,450 Long COVID cases and 1,093,995 population controls from 24 studies across 16 countries. We identified the first genome-wide significant association for Long COVID at the *FOXP4* locus. *FOXP4* has been previously associated with COVID-19 severity^6^, lung function^8^, and cancers^9^, suggesting a broader role for lung function in the pathophysiology of Long COVID. While we identify COVID-19 severity as a causal risk factor for Long COVID, the impact of the genetic risk factor located in the *FOXP4* locus could not be solely explained by its association to severe COVID-19. Our findings further support the role of pulmonary dysfunction and COVID-19 severity in the development of Long COVID.

## Introduction

The COVID-19 pandemic has led to the recognition of a new condition known as post-acute sequelae of COVID-19 (PASC), post COVID-19 condition, or Long COVID. The current World Health Organization definition includes any symptoms that present after COVID-19 and persist after three months^10^. Common symptoms include fatigue, pulmonary dysfunction, muscle and chest pain, dysautonomia and cognitive disturbances^11–15^. The incidence of Long COVID varies widely, with estimates ranging from 10% to 70%^5^. Long COVID is more common in individuals who have been hospitalized or treated at the intensive care unit due to COVID-19, suggesting that COVID-19 severity could contribute to the risk of Long COVID^5, 16^. However, Long COVID can also occur in those with initially mild COVID-19 symptoms^17^. Potential mechanisms for Long COVID include persistent COVID-19 infection, autoimmunity, reactivation of latent pathogens such as chickenpox or Epstein Barr virus, disrupted blood clotting, and dysregulation of the autonomic nervous system^5^.

The COVID-19 Host Genetics Initiative (COVID-19 HGI) (https://www.covid19hg.org) was launched to investigate the role of host genetics in COVID-19 and its various clinical subtypes^18, 19^. The COVID-19 HGI has identified 51 distinct genome-wide significant loci associated with COVID-19 critical illness, hospitalization and SARS-CoV-2 reported infection. These variants largely implicate canonical pathways involved in viral entry, mucosal airway defence, and type I interferon response^6, 7, 20, 21^.

To better understand the underlying causes of Long COVID, we conducted the first genome-wide association study (GWAS) specifically focused on Long COVID. Our study includes data from 24 studies conducted in 16 countries, totalling 6,450 individuals diagnosed with Long COVID and 1,093,995 controls (**Fig. 1**).

**Fig. 1.**
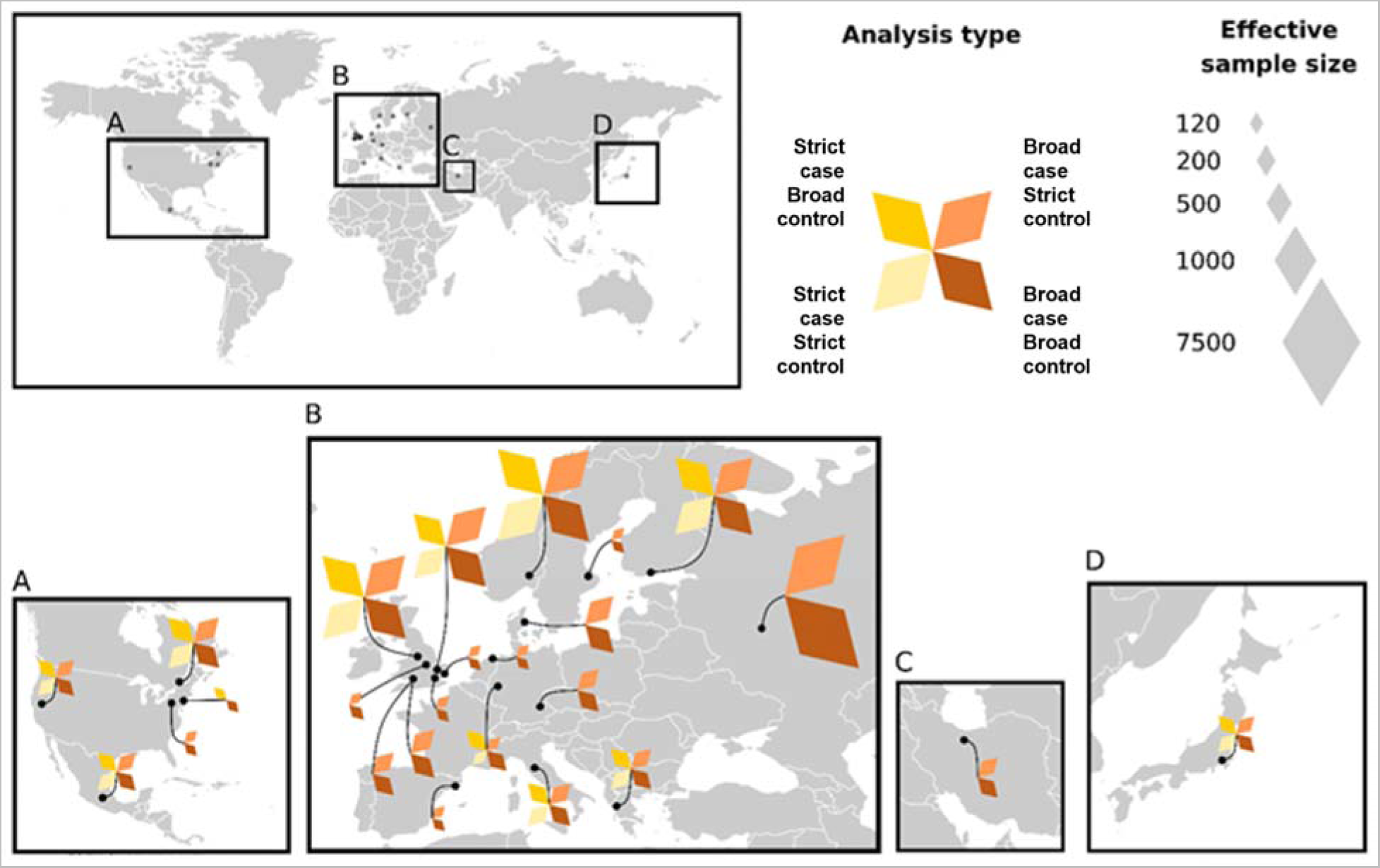
Geographic overview of studies contributing to analysis of Long COVID. The 24 studies contributing to the Long COVID HGI Data Freeze 4 GWAS meta-analyses. Each colour represents a meta-analysis with specific case and control definitions. Strict case definition = Long COVID after test-verified SARS-CoV2 infection, broad case definition = Long COVID after any SARS-CoV-2 infection. Strict control definition = individuals that had SARS-CoV-2 but did not develop Long COVID, broad control definition = population control i.e. all individuals in each study that did not meet the Long COVID criteria. Effective sample sizes are shown as the size of each diamond shape. For more detailed sample sizes, see **Extended Table S1**.

## Results

### Genetic variants in the FOXP4 locus are associated with Long COVID

We analysed 24 independent GWAS of Long COVID and computed four GWAS meta-analyses based on two case definitions and two control definitions. A strict Long COVID case definition required having an earlier test-verified SARS-CoV-2 infection (strict case definition), while a broader Long COVID case definition also included self-reported or clinician-diagnosed SARS-CoV-2 infection (broad case definition). The broad definition included all of the contributing studies whereas the strict definition was met by 11 studies (**Extended Table S1**). Controls were either population controls, i.e., genetically ancestry-matched samples without known Long COVID (broad control definition), or people that had recovered from SARS-CoV-2 infection without Long COVID (strict control definition) (**Fig. 1, Extended Table S2**). Data was obtained from altogether 16 countries, representing populations from 6 genetic ancestries, and each meta-analysis combined data across ancestries. The most common symptoms in the questionnaire-based studies with available symptom information were fatigue, shortness of breath, and problems with memory and concentration. However, there was some heterogeneity in the frequency of symptoms (**Extended Fig. S1**).

The GWAS meta-analysis using the strict case definition (N = 3,018) and the broad control definition (N = 994,582) from 11 studies identified a genome-wide significant association within the *FOXP4* locus (chr6:41,515,652G>C, GRCh38, rs9367106, as the lead variant; P = 1.8×10^-10^, **Fig. 2, Table S3**). The C allele at rs9367106 was associated with an increased risk of Long COVID (OR = 1.63, 95% confidence interval (CI): 1.40-1.89, risk allele frequency = 4.2%).

**Fig. 2.**
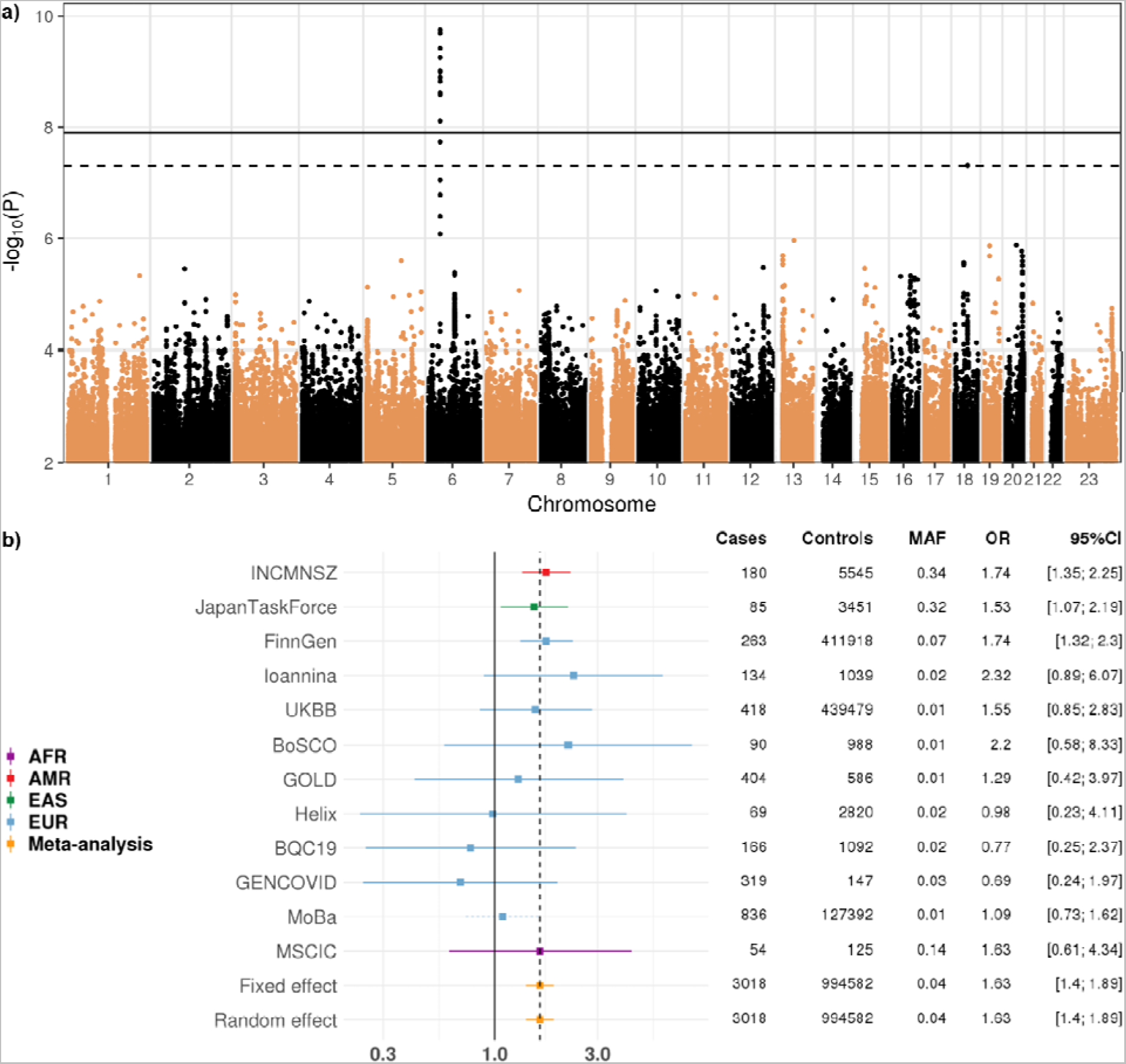
Meta-analysis of 11 GWAS studies of Long COVID shows an association at the *FOXP4* locus. a) Manhattan plot of Long COVID after test-verified SARS-CoV-2 infection (strict case definition, N = 3,018) compared to all other individuals in each data set (population controls, broad control definition, N = 994,582). A genome-wide significant association with Long COVID was found in the chromosome 6, upstream of the *FOXP4* gene (chr6:41515652:G:C, GRCh38, rs9367106, as the lead variant; P = 1.76×10^-10^, Bonferroni P = 7.06×10^-10^, increased risk with the C allele, OR = 1.63, 95% CI: 1.40-1.89). Horizontal lines indicate genome-wide significant thresholds before (PlJ<lJ5×10^−8^, dashed line) and after (1.25×10^−8^) Bonferroni correction over the four Long COVID meta-analyses. b) Chromosome 6 lead variant across the contributing studies and ancestries GWAS meta-analyses of Long COVID with strict case definition and broad control definition. Lead variant rs9367106 (solid line) and if missing, imputed by the variant with the highest linkage disequilibrium (LD) with the lead variant for illustrative purpose, i.e. rs12660421 (r = 0.98 in European in 1000G+HGDP samples^49^, dotted lines). For the imputed variants, beta was weighted by multiplying by the LD correlation coefficient (r = 0.98).

We observed an association, albeit not genome-wide significant, with rs9367106-C and Long COVID also in all other three meta-analyses, including our largest meta-analysis with the broad case definition (N = 6,450) and the broad control definition (N = 1,093,995) from 24 studies (OR = 1.34, 95% CI: 1.20-1.49, P = 1.1×10^-7^, **Extended Fig. S2, S3**). Analyses with the strict case definition (N = 2,975) and strict control definition (N = 37,935) (OR = 1.30, 95% CI: 1.09-1.56, P = 3.8×10^-3^), or the broad case definition (N = 6,407) and strict control definition (N = 46,208) (OR = 1.16, 95% CI: 1.02-1.32, P = 0.023), further supported our finding (**Extended Fig. S3**).

To examine the consistency of the *FOXP4* signal across the contributing studies, we investigated the effect in each study. As a subset of studies lacked data for the lead variant, we used the available variant in highest linkage disequilibrium (LD) as a proxy (**Fig. 2b**). Genetic variants in the meta-analysis had varying statistical power due to missingness, due to genotyping and imputation quality, and due to differences in allele frequency differences between populations. Therefore, the genetic variant that was present in the majority of the studies was the most significant variant, not because it is the causal variant but because it had the best statistical power. Moreover, variants in high LD with the same, or similar, effect could display different association strengths because of the different statistical power across the variants. We therefore examined the effect size of variants within 30 kb around the most significant variant (rs9367106), including variants even with weak LD with the lead variant (r^2^ > 0.01 in individuals of Europeans in the Human Genome Diversity Project^22^ and 1000 Genome Project^23, 24^ and effective sample size at least one third the sample size of the lead variant. Through this analysis, we identified a haplotype spanning the genomic region chr6:41,512,355-41,537,458 (Genome Reference Consortium Human Build 38, GRCh38), located upstream of *FOXP4* gene, for which variants had a similar effect size to the lead variant (**Fig. 3b**) and P values less than 5×10^-7^. This analysis identified 15 variants (**Extended Table S4**). Relying on LD in the 1000 Genomes project among Europeans, we found 18 variants co-segregating with the lead variant (r^2^ > 0.5, **Extended Table S5**). 9 variants overlapped between these two analyses. None of the variants identified were protein coding.

**Fig. 3.**
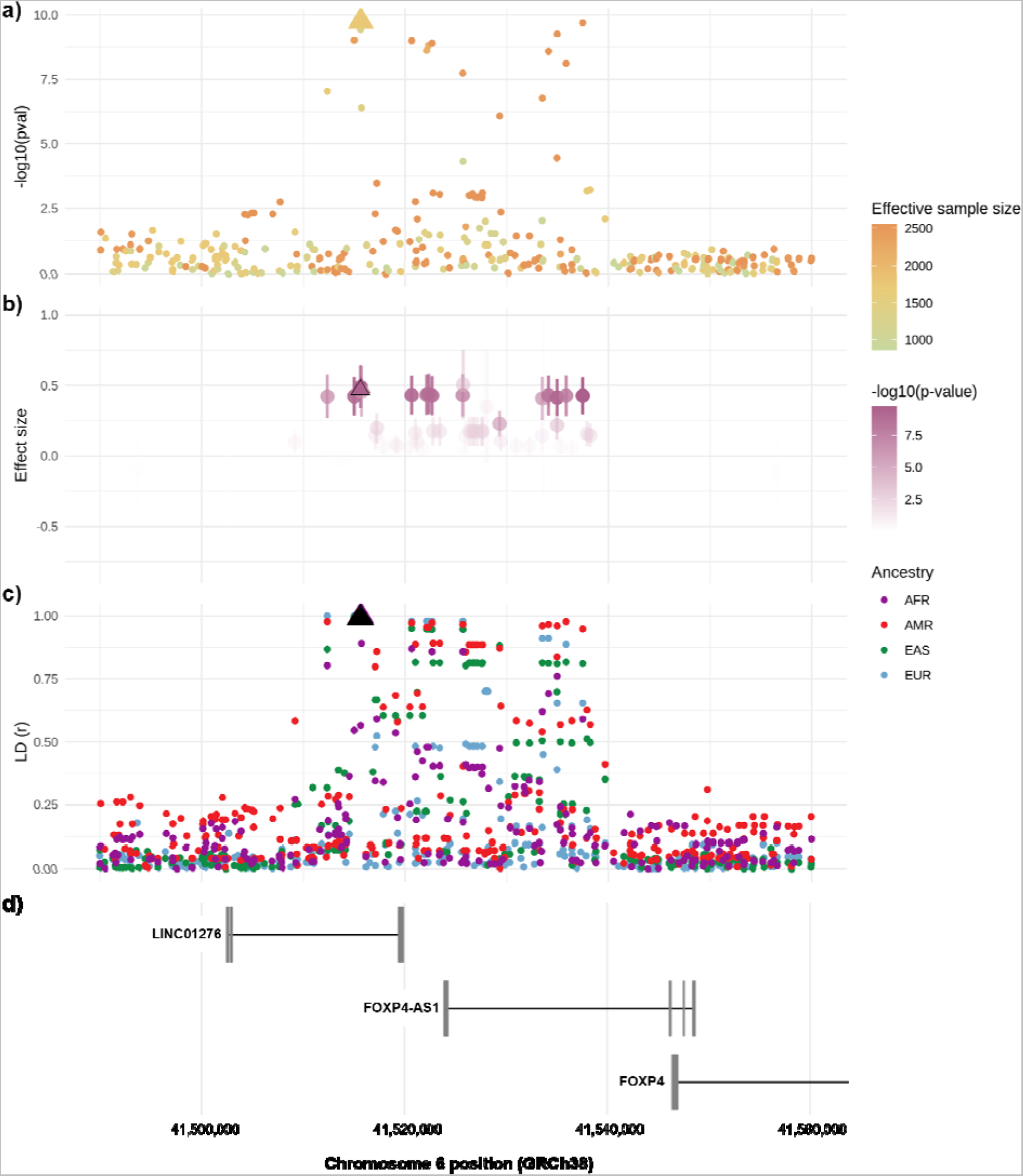
The chromosome 6 region (chr6:41,490,001-41,560,000 (70 kb); *FOXP4* locus) in the Long COVID GWAS meta-analysis. Long COVID meta-analysis with strict case and broad control definition (see **Fig. 2**). X-axis shows the position on chromosome 6 (Genome Reference Consortium Human Build 38). The Long COVID lead variant (rs9367106) depicted with a triangle in each plot. **a)** Locus zoom plot with each variant coloured by effective sample size and showing statistical significance on y-axis. **b)** Each variant coloured by statistical significance (-log_10_(P value)) and showing effect sizes (beta ± standard error). **c)** Each variant coloured by ancestry and showing linkage disequilibrium (r) with our lead variant on y-axis. AFR, African; AMR, Admixed American; EAS, East Asian; EUR, European. **d)** Ensembl genes in the region (*FOXP4* not fully shown) (www.ensembl.org)^50^.

### Frequency of Long COVID variants at FOXP4 varies across ancestries

The allele frequency of rs9367106-C at the *FOXP4* locus varied greatly among the different study populations, with frequencies ranging from 1.6% in studies with non-Finnish Europeans to higher frequencies such as 7.1% in Finnish, 19% in admixed Americans, and 36% in East Asians (**Extended Fig. S4**). Most of the studies included in our analysis had individuals of primarily European descent (**Extended Fig. S5**). Despite smaller sample sizes, we observed significant associations for the *FOXP4* variant in the studies with admixed American, East Asian, and Finnish ancestries (**Fig. 2b**), owing to the higher allele frequency, and thus larger statistical power to detect an association with the rs9367106 variant in these cohorts.

### FOXP4 risk variants increase the expression of FOXP4 in the lung and is associated with COVID-19 severity

The genomic region (+/-100 kilobases) surrounding the lead variant associated with Long COVID contains four genes (*FOXP4*, *FOXP4-AS1*, *LINC01276*, *MIR4641*). Since no variant in LD with the lead variant is coding, we investigated if any of these variants were associated with differential expression of any of the surrounding genes within a 100 kb window. We used rs12660421 as a proxy (rs12660421-A allele is correlated with the Long COVID risk allele rs9367106-C, r^2^ = 0.97 among European ancestry individuals), given that the lead variant was not included in the GTEx dataset V8, and analysed differential gene expression across all tissues included in the dataset. We found that rs12660421-A is associated with an increase in *FOXP4* expression in the lung (P = 5.3×10^-9^, normalized effect size (NES) = 0.56) and in the hypothalamus (P = 2.6×10^-6^, NES = 1.4) (**Fig. 4, Extended Fig. S6**). None of the other genes demonstrated any differential expression regarding the Long COVID-associated haplotype. *FOXP4* is a transcription factor gene which has a broad tissue expression pattern and is expressed in nearly all tissues, with the highest expression in the cervix, the thyroid, the vasculature, the stomach, and the testis^25^. The expression also spans a broad set of cell types, including endothelial lung cells, immune cells, and myocytes^26^. A colocalization analysis (**Methods**) suggested that the association signal of Long COVID is the same signal that associates with the differential expression of *FOXP4* in the lung (posterior probability = 0.91) (**Extended Fig. S7a,b**, **Extended Table S6**).

**Fig. 4.**
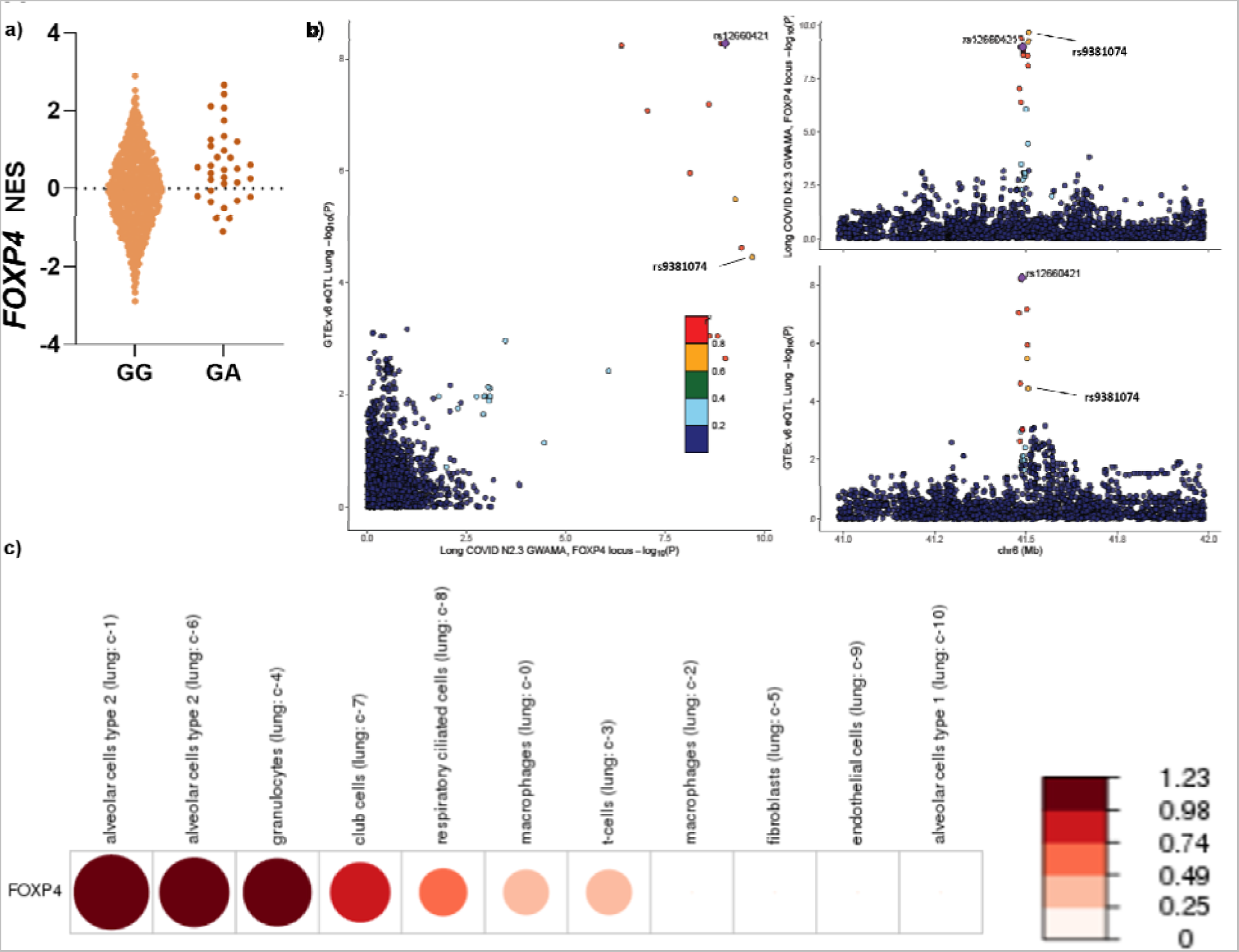
*FOXP4* expression in the lung. **a)** The lead variant rs9367106 was not found in the GTEx dataset, but a proxy variant (rs12660421, chr6:41520640) in high LD (r^2^=0.97, rs12660421-A allele is correlated with the Long COVID risk allele rs9367106-C) showed a significant expression quantitative trait locus (eQTL), increasing *FOXP4* expression in the lung (P = 5.3×10^-9^, normalized effect size (NES) = 0.56, https://gtexportal.org/home/snp/rs12660421). For other tissues, see multi-tissue eQTL plot in the Supplemental information (**Extended Fig. S6**). **b)** Colocalization analysis using eQTL data from GTEx v8 tissue type and Long COVID association data. Plots illustrate -log_10_ P value for Long COVID (x-axis) and for *FOXP4* expression in the Lung (y-axis), regional association of the *FOXP4* locus variants with Long COVID (top right), and regional association of the *FOXP4* variants with RNA expression measured in the Lung in GTEx (bottom right). Variants are coloured by 1000 Genomes European-ancestry LD r^2^ with the lead variant (rs12660421) for *FOXP4* expression in lung tissue. (The most significant Long COVID variant overlapping the GTEx v8 dataset (rs9381074) also annotated.) **c)** Human Protein Atlas RNA single cell type tissue cluster data (transcript expression levels summarized per gene and cluster) of lung (GSE130148) showing *FOXP4* expression in unaffected individuals. The values were visualized using log10(protein-transcripts per million [pTPM]) values. Each c-X annotation is taken from the clustering results performed in Human Protein Atlas.

Furthermore, variants in this region have also been identified as risk factors for hospitalization due to COVID-19 in the COVID HGI meta-analyses^6^ and in Biobank Japan (**Extended Fig. S8, Extended Table S7**). Our colocalization analysis demonstrated the *FOXP4* risk haplotype identified here as the same haplotype identified for COVID-19 severity (posterior probability > 0.97) (**Extended Fig. S7e,f**, **Extended Table S6**).

### Single cell sequencing data supports FOXP4 expression in alveolar and immune cells in the lung

As lung tissue consists of several cell types, we wanted to elucidate the relevant cells that express *FOXP4* and may contribute to Long COVID. To understand the role of *FOXP4* in healthy lung before SARS-CoV-2 infection, we analysed single cell sequencing data from the Tabula Sapiens (data available through the Human Protein Atlas; https://www.proteinatlas.org/), a previously published atlas of single cell sequencing data in healthy individuals free of COVID-19^27^. We observed the highest expression of *FOXP4* in type 2 alveolar cells (**Fig. 4**), a cell type that is capable of mounting robust innate immune responses, thus participating in the immune regulation in the lung^28^. Furthermore, type 2 alveolar cells secrete surfactant, keep the alveoli free from fluid, and serve as progenitor cells repopulating damaged epithelium after injury^29^. In addition, we observed nearly equally high expression of *FOXP4* in granulocytes that similarly participate in regulation of innate immune responses. Overall, the findings suggest a possible role of both immune and alveolar cells in lung in Long COVID.

### The Long COVID FOXP4 variants are located at active chromatin in the lung

To understand the regulatory effects behind the variant association, we utilized the data from the Regulome database^30, 31^, ENCODE^32^, and VannoPortal^33^. We discovered that while the majority of the Long COVID variants had active enhancer or transcription factor binding in a few ENCODE experiments, we identified four variants of interest with possible additional functional consequences (**Extended Tables S8, S9**). These variants had direct evidence of transcription factor binding based on Chip sequencing experiments. rs2894439 located at the beginning of the risk haplotype was bound by eight transcription factors, including POLR2A and EP300. rs7741164 and rs55889968 were both bound by six transcription factors, including EP300 and FOXA1. And finally, one variant (rs9381074) was directly located on a region that had DNA methylation marks across multiple tissues, including immune and lung cells (H3K27me3 and H3K4me1, H3K4me3, H3K27ac, H3K4me2, H3K4me3), and had evidence of transcriptional activity from 49 different transcription factors, of which we saw the most consistent direct binding of FOXA1 across 55 experiments. Furthermore, we downloaded DNase sequencing data from the ENCODE project and observed that rs9381074 was directly positioned on a DNase hypersensitivity site in the lung (see **Supplementary Methods** for accession numbers).

### The Long COVID FOXP4 variant is associated with lung cancer

To further understand the genetic variant that increases the risk of Long COVID, we examined whether the *FOXP4* variant was also associated with any other diseases. Specifically, we focused on Biobank Japan^34^, as the Long COVID risk allele frequency is highest in East Asia. Phenome-wide association study between rs9367106 and all phenotypes in Biobank Japan (N = 262) revealed that Long COVID risk allele was associated with lung cancer (P = 1.2×10^-6^, Bonferroni P = 3.1×10^-4^, OR = 1.13, 95% CI = 1.07-1.18) (**Extended Fig. S8, Extended Table S7**). Furthermore, the Long COVID risk allele is in LD with the known risk variants for non-small cell lung carcinoma in Chinese and European populations^35^ (rs1853837, r^2^ = 0.88 in East Asian^36^) and for lung cancer in never-smoking Asian women^37^ (rs7741164, r^2^ = 0.98 in East Asian^36^). Colocalization analysis supported that the associations in this locus (within 500 kb of rs9367106) for Long COVID and lung cancer shared the same genetic signal (colocalization posterior probability = 0.98, **Extended Fig. S7c,d**).

### Long COVID and other phenotypes

We investigated the relationship between Long COVID and cardiometabolic, behavioural, and psychiatric traits^7^ (**Fig. 5, Extended Table S10**). We found positive genetic correlations between Long COVID and insomnia symptoms, depression, risk tolerance, asthma, diabetes, and SARS-CoV-2 infection, while we saw negative correlations with red and white blood cell counts (**Fig. 5a**). However, identified correlations were only nominally significant without multiple testing correction (P < 0.05; **Extended Table S11**). The estimated heritability of Long COVID was h^2^ = 0.023 in the meta-analysis using the strict case and control definitions.

**Fig. 5.**
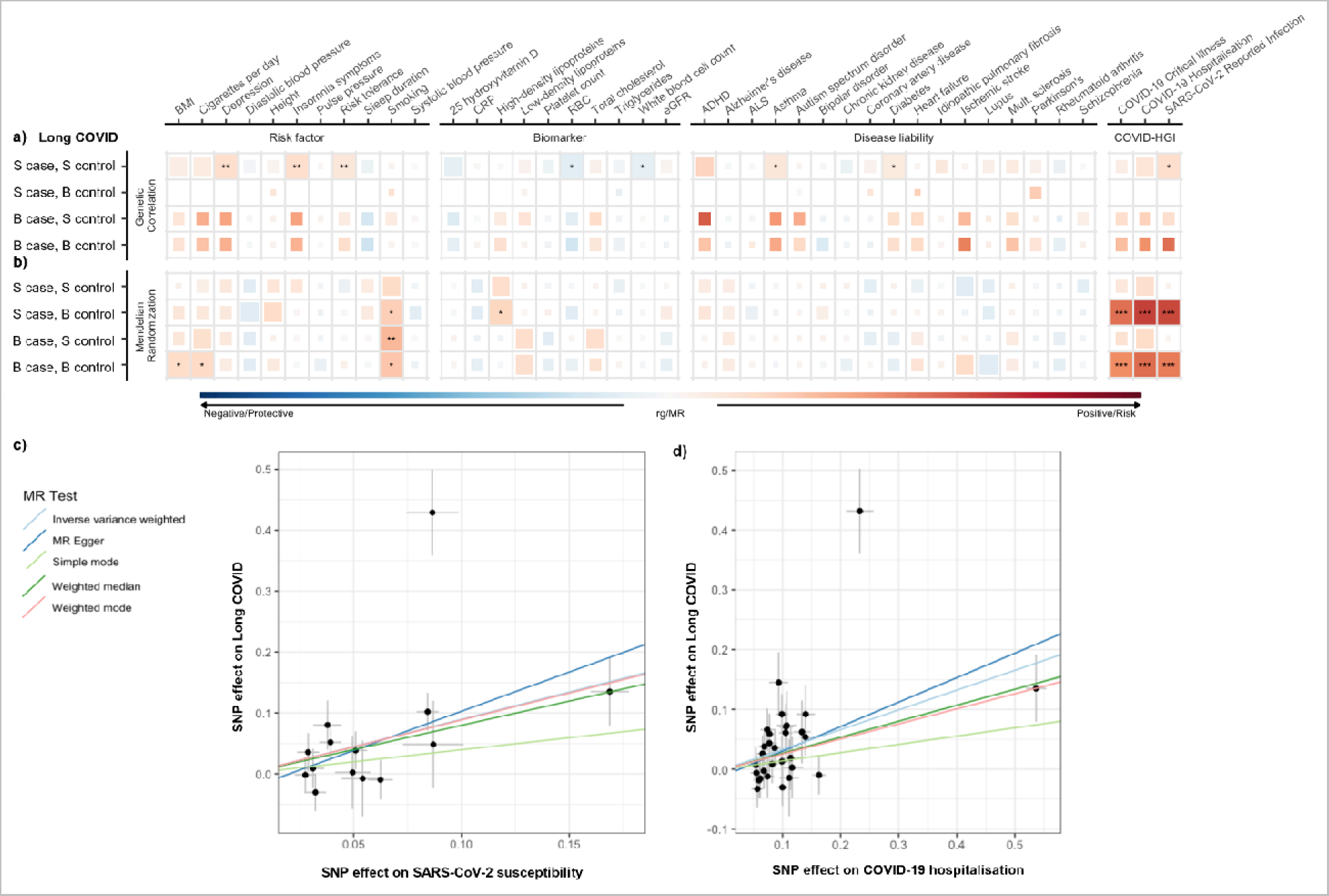
Genetic correlations and Mendelian randomization causal estimates between Long COVID and potential risk factors, biomarkers and diseases. **a)** Linkage disequilibrium score regression (LDSC, upper panel; **Extended Table S11**) and **b)** inverse variance-weighted Mendelian randomization (MR, bottom panel; **Extended Table S12**) were used for calculating two-sided P values. Size of each coloured square corresponds to statistical significance (P values < 0.01 [**] full-sized square, <0.05 [*] full-sized square, <0.1 large square, <0.5 medium square, and >0.5 small square; not corrected for multiple comparisons). BMI, body mass index; CRP, C-reactive protein; eGFR, estimated glomerular filtration rate; ADHD, attention-deficit hyperactivity disorder. **b)** MR scatter plot with effect sizes (beta±SE) of each variant on COVID-19 susceptibility (reported SARS-CoV-2 infection) as exposure and Long COVID (strict case, broad control definition) as outcome (P IVW = 2.4×10^−3^, pleiotropy P = 0.47; **Extended Table S13**). **c)** Similarly, MR with COVID-19 hospitalization as exposure and Long COVID as outcome (P IVW = 7.5×10^−5^, pleiotropy P = 0.55; **Extended Table S13**).

We used Mendelian randomization (MR) to estimate potential risk factors by analysing the same traits mentioned above. Genetically predicted earlier smoking initiation (P = 0.022), more cigarettes consumed per day (P = 0.046), higher levels of high-density lipoproteins (P = 0.029), and higher body-mass index (P = 0.046) were nominally significant causal risk factors of Long COVID (**Fig. 5b, Extended Table S12**). However, none of these associations survived correction for multiple comparisons.

### The FOXP4 signals cannot be explained simply by severity of acute COVID-19

Earlier research has suggested that COVID-19 severity may be a risk factor for Long COVID^3, 16, 38, 39^. We investigated the relationship between COVID-19 hospitalization and Long COVID by performing a two-sample MR (**Extended Table S13**). In terms of causality, we caution that COVID-19 hospitalization as causal exposure is difficult to interpret because both Long COVID and COVID-19 hospitalization are two outcomes of the same underlying infection. Nevertheless, the relationship between the effect size for Long COVID versus the effect size for COVID-19 severity can shed some light on the role of COVID-19 severity in Long COVID. To perform two-sample MR without overlapping samples, we have excluded the studies that contributed to the current Long COVID freeze 4 and re-run a meta-analysis of COVID-19 susceptibility and hospitalization of the remaining cohorts in the COVID-19 HGI. We observed a causal relationship of susceptibility and hospitalization on Long COVID (inverse-variance weighted MR, P ≤ 2.4×10-3, for the strict case and the broad control definition) with no evidence of pleiotropy (MR Egger intercept P ≥ 0.47) (**Fig. 5c,d, Extended Table S13**). Nevertheless, the Wald ratio of Long COVID to COVID-19 hospitalization for the *FOXP4* variant is 1.97 (95% CI: 1.36-2.57), which is significantly greater than the slope of the MR-estimated relationship between COVID-19 hospitalization and Long COVID (0.35, 95% CI: 0.12-0.57). The same phenomenon was seen when comparing with susceptibility (**5c**). Thus, the *FOXP4* signal demonstrates a stronger association with Long COVID than expected, meaning that it cannot simply be explained by its association with either susceptibility or severity alone (**Fig. 5c,d**). A recent systematic review of epidemiological data found a positive association between COVID-19 hospitalization and Long COVID with a relationship on a log-odds scale of 0.91 (95% CI: 0.68 - 1.14)^40^. Even assuming this stronger relationship between COVID-19 hospitalization and Long COVID, the observed effect of the *FOXP4* variant on Long COVID still exceeds what would be expected based on the association with severity alone.

## Discussion

In this study, we aimed to understand the host genetic factors that contribute to Long COVID, using data from 24 studies across 16 countries. Our analysis identified genetic variants within the *FOXP4* locus as a risk factor for Long COVID. The *FOXP4* gene is expressed in the lung and the genetic variants associated with Long COVID are also associated with differential expression of *FOXP4* and with lung cancer and COVID-19 severity. Additionally, using MR, we characterized COVID-19 severity as a causal risk factor for Long COVID. Overall, our findings provide genomic evidence consistent with previous epidemiological and clinical reports of Long COVID, indicating that Long COVID, similarly to other post-viral conditions, is a heterogeneous disease entity where likely both individual genetic variants and the environmental risk factors contribute to disease risk.

Our analysis revealed a connection between Long COVID and pulmonary endpoints through both individual variants at *FOXP4*, a transcription factor-coding gene previously linked to lung cancer, and MR analysis identifying smoking and COVID-19 severity as risk factors. Furthermore, expression analysis of the lung, and cell type-specific single-cell sequencing analysis, showed *FOXP4* expression in both alveolar cell types and immune cells of the lung. *FOXP4* belongs to the subfamily P of the forkhead box transcription factor family genes and is expressed in various tissues, including the lungs and gut^41, 42^. Moreover, it is highly expressed in mucus-secreting cells of the stomach and intestines^43^, as well as in naïve B, natural killer, and memory T-reg cells^44^, and required for normal T-cell memory function following infection^45^. *FOXP1/2/4* are also required for promoting lung endoderm development by repressing expression of non-pulmonary transcription factors^46^, and the loss of *FOXP1/4* adversely affects the airway epithelial regeneration^8^. Furthermore, *FOXP4* has been implicated in airway fibrosis^47^ and promotion of lung cancer growth and invasion^48^. We find that the haplotype associated with Long COVID is also associated with lung cancer in Biobank Japan^34^. These observations together with the present study may suggest that the connection between *FOXP4* and Long COVID may be rooted in both lung function and immunology. Furthermore, the observation of *FOXP4* expression in both alveolar and immune cells in the lung, and the association with severe COVID-19 and pulmonary diseases such as cancer, suggest that *FOXP4* may participate in local immune responses in the lung.

We also discovered a causal relationship from COVID-19 infection to Long COVID, as expected, and an additional causal risk from severe, hospital treatment-requiring COVID-19 to Long COVID. This finding is in agreement with earlier epidemiological studies where higher prevalence of Long COVID was seen among individuals that had severe acute COVID-19 infection^3, 16, 38, 39^. The observation between COVID-19 severity and Long COVID raises an interesting question: When SARS-CoV-2 infection is required for COVID-19, and severe COVID-19, are all genetic variants that increase COVID-19 susceptibility or severity equally large risk factors for Long COVID? In the current study, we aimed to answer this question through examining variant effect sizes between SARS-CoV-2 infection susceptibility, COVID-19 severity, and Long COVID. We discovered that the majority of variants affected only SARS-CoV-2 susceptibility or COVID-19 severity. In contrast, the *FOXP4* variants had higher effect size for Long COVID than expected, suggesting an independent role of *FOXP4* for Long COVID that was not observed with overall COVID-19 severity variants. Such observation offers clues on biological mechanisms, such as *FOXP4* affecting pulmonary function and immunity, which then contribute to the development of Long COVID. Overall, our study elucidates genetic risk factors for Long COVID, the relationship between Long COVID and severe COVID-19, and finally possible mechanisms of how *FOXP4* contributes to the risk of Long COVID. Future studies and iterations of this work will likely grow the number of observed genetic variants and further clarify the biological mechanisms underlying Long COVID.

We recognize that the symptomatology of Long COVID is variable and includes in addition to lung symptoms, also other symptom domains such as fatigue and cognitive deficits^3–5^. In addition, the long-term effects of COVID-19 are still being studied, and more research is needed to understand the full extent of the long-term damage caused by SARS-CoV-2 and Long COVID disease. We also recognize that the Long COVID diagnosis is still evolving. Nevertheless, our study provides direct genetic evidence that lung pathophysiology can play an integral part in the development of Long COVID.

## Methods

### Contributing studies

A total of 24 studies contributed to the analysis, with a total sample size of 6,450 Long COVID cases with 46,208 COVID-19 positive controls and 1,093,955 population controls. Participants provided informed consent to participate in the respective study, with recruitment and ethics following study-specific protocols approved by their respective Institutional Review Boards (Details are provided in **Extended Table S2**). The effective sample sizes for each study shown in **Fig. 1** were calculated for display using the formula: (4 × N_case × N_control)/(N_case + N_control). The Long COVID Host Genetics Initiative is a global and ongoing collaboration, open to all studies around the world that have data to run Long COVID GWAS using our phenotypic criteria described below.

### Phenotype definitions

We used the following criteria for assigning case control status for Long COVID aligning with the World Health Organization guidelines^10^ (**Supplementary Methods**). Study participants were defined as Long COVID cases if, at least three months since SARS-CoV-2 infection or COVID-19 onset, they met any of the following criteria:

1. presence of one or more self-reported COVID-19 symptoms that cannot be explained by an alternative diagnosis
2. report of ongoing significant impact on day-to-day activities
3. any diagnosis codes of Long COVID (e.g. Post COVID-19 condition, ICD-10 code U09(.9)) Criteria 1 and 2 were applied only to questionnaire-based cohorts, whereas 3 was used in studies with electronic health records (EHR). Detailed phenotyping criteria and diagnosis codes of each study are provided in **Extended Table S2**.

We used two Long COVID case definitions, a strict definition requiring a test-verified SARS-CoV-2 infection and a broad definition including self-reported or clinician-diagnosed SARS-CoV-2 infection (any Long COVID).

We applied two control definitions. First, we used population controls, i.e. everybody that is not a case. Population controls were genetic ancestry-matched individuals who were not defined as Long COVID cases using the above-mentioned questionnaire or EHR-based definition. In the second analysis, we compared Long COVID cases to individuals who had had SARS-CoV-2 infection but who did not meet the criteria of Long COVID, i.e. had fully recovered within 3 months from the infection.

We used in total four different case-control definitions to generate four GWASs as below;

1. Long COVID cases after test-verified SARS-CoV-2 infection vs population controls (the strict case definition vs the broad control definition)
2. Long COVID within test-verified SARS-CoV-2 infection (the strict case definition vs the strict control definition)
3. Any Long COVID cases vs population controls (the broad case definition vs the broad control definition)
4. Long COVID within any SARS-CoV-2 infection (the broad case definition vs the strict control definition)

### Genome-wide association studies

We largely applied the GWAS analysis plans used in the COVID-19 HGI^6^. Each study performed their own sample collection, genotyping, genotype and sample quality control (QC), imputation and association analyses independently, according to our central analysis plan (https://docs.google.com/document/d/1XRQgDOEp62TbWaqLYi1RAk1OHVP5T3XZqfs_6PoPt_k), before submitting the results for meta-analysis (Details are provided in **Extended Table S2**). We recommended that GWAS were run using REGENIE^51^ on chromosomes 1–22 and X, though some studies used SAIGE^52^ or PLINK^53^. The minimum set of covariates to be included at runtime were age, age^2^, sex, age × sex and the first 10 genetic principal components. We advised studies to include any additional study-specific covariates where needed, such as those related to genotype batches or other demographic and technical factors that could lead to stratification within the cohort. Studies performing the GWAS using a software that does not account for sample relatedness (such as PLINK) were advised to exclude related individuals.

### GWAS meta-analyses

The meta-analysis pipeline was also adopted from the COVID-19 HGI flagship paper [https://www.nature.com/articles/s41586-021-03767-x]. The code is available at LongCOVID HGI GitHub (https://github.com/long-covid-hg/META_ANALYSIS/), and is a modified version of the pipeline developed for the COVID-19 HGI (https://github.com/covid19-hg/META_ANALYSIS). To ensure that individual study results did not suffer from excessive inflation, deflation and false positives, we manually investigated plots of the reported allele frequencies against aggregated gnomAD v3.0 ^49^ allele frequencies in the same population. We also evaluated whether the association standard errors were excessively small, given the calculated effective sample size, to identify studies deviating from the expected trend. Where these issues were detected, the studies were contacted to reperform the association analysis, if needed, and resubmit their results.

Prior to the meta-analysis itself, the summary statistics were standardized, filtered (excluding variants with allele frequency <0.1% or imputation INFO score <0.6), lifted over to reference genome build GRCh38 (in studies imputed to GRCh37), and harmonized to gnomAD v3.0 through matching by chromosome, position and alleles (**Supplementary Methods**).

The meta-analysis was performed using a fixed-effects inverse-variance weighted (IVW) method on variants that were present in at least two studies contributing to the specific phenotype being analysed. To assess if one study was primarily driving any associations, we simultaneously ran a leave-most-significant-study-out (LMSSO) meta-analysis for each variant (based on the variant’s study-level P value). Heterogeneity between studies were estimated using Cochran’s Q-test^54^. Each set of meta-analysis results were then filtered to exclude variants whose total effective sample size (in the non-LMSSO analysis) was less than 1/3 of the total effective sample size of all studies contributing to that meta-analysis. We report significant loci that pass the genome-wide significance threshold (P ≤ 5×10^-8^ / 4 = 1.25×10^-8^) accounting for the number of GWAS meta-analyses we performed.

### Principal component projection

In a similar fashion to the COVID-19 HGI, we asked each study to project their cohort onto a multi-ethnic genetic principal component space (**Extended Fig. S5**), by providing studies with pre-computed PC loadings and reference allele frequencies from unrelated samples from the 1000 Genomes Project^23, 24^ and the Human Genome Diversity Project. The loadings and frequencies were generated for a set of 117,221 autosomal, common (MAF ≥ 0.1%) and LD-pruned (r2 < 0.8; 500-kb window) single-nucleotide polymorphisms (SNPs) that would be available in the imputed data of most studies. Access to the projecting and plotting scripts was made available to the studies at https://github.com/long-covid-hg/pca_projection.

### eQTL, PheWAS and colocalization

For the single (Bonferroni-corrected) genome-wide significant lead variant, rs9367106, we used the GTEx portal (https://gtexportal.org/)^26^ to understand if this variant had any tissue-specific effects on gene expression. As rs9367106 was not available in the GTEx database, we first identified a proxy variant, rs12660421 (r^2^ = 0.90) using all individuals from the 1000 Genomes Project^23^ and then performed a lookup in the portal’s GTEx v8 dataset^25^.

To identify other phenotypes associated with rs9367106, we used the Biobank Japan PheWeb portal (https://pheweb.jp/)^9^ to perform a phenome-wide association analysis, as the minor allele frequency of rs9367106 is highest in East Asia. To assess whether the *FOXP4* association is shared between Long COVID, and tissue-specific eQTLs, lung cancer, and COVID-19 hospitalization, we extracted a 1Mb region centred on rs9367107 (chr6:41,015,652-42,015,652) from the lung cancer and COVID-19 hospitalization summary statistics and the GTEx v8 data and performed colocalization analyses using the R package coloc (v5.1.0.1)^55, 56^ in R v4.2.2. Colocalization locus zoom plots were created using the *LocusCompareR* R package v1.0.0^57^ with LD r^2^ estimated using 1000 Genomes European-ancestry individuals^23, 24^.

### Genetic correlation and Mendelian Randomization

We assessed the genetic overlap and causal associations between Long COVID outcomes and the same set of risk factors, biomarkers and diseases liabilities as in the COVID-19 HGI flagship paper^6^. Additionally, we tested the overlap and causal impact of COVID-19 susceptibility and hospitalization risk. Genetic correlations were assessed using Linkage Disequilibrium Score Regression (LDSC) v1.0.1 ^58, 59^. Where there were sufficient genome-wide significant variants, the causal impact was tested in a two-sample Mendelian Randomization framework using the TwoSampleMR (v0.5.6) R package^60^ with R v4.0.3. To avoid sample overlap between exposure GWASs (here COVID-19 hospitalization and SARS-CoV-2 reported infection) and outcome GWASs (here Long COVID phenotypes), we performed meta-analyses of COVID-19 hospitalization and SARS-CoV-2 reported infection using data freeze 7 of the COVID-19 HGI by excluding studies that participated in the Long COVID (freeze 4) effort. Independent significant exposure variants with p ≤ 5×10^-8^ were identified by LD-clumping the full set of summary statistics using an LD r^2^ threshold of 0.001 (based on the 1000 Genomes European-ancestry reference samples^23^) and a 10-Mb clumping window. For each exposure-outcome pair, these variants were then harmonized to remove variants with mismatched alleles and ambiguous palindromic variants (MAF >45%). Fixed-effects Inverse Variance Weighted meta-analysis was used as the primary MR methods, with MR-Egger, Weighted Median Estimator, Weighted Mode Based Estimator, MR-PRESSO used in sensitivity analyses. Heterogeneity was assessed using the MR-PRESSO global test and pleiotropy using the MR-Egger intercept. The genetic correlation and Mendelian Randomization analysis were implemented as a Snakemake Workflow made available at https://github.com/marcoralab/MRcovid.

Summaries of the exposure GWAS are provided in **Extended Table S10** and the association statistics for all exposure variants are provided in **Extended Table S14**.

## Data availability

We have made the results of these GWAS meta-analyses publicly available for variants passing post-meta-analysis filtering for minor allele frequency >=1% and effective sample size >1/3 of the maximum effective sample size for each meta-analysis. These can be accessed at LocusZoom, where the associations can be visually explored and the summary statistics exported for further scientific discovery^61^.

Strict case definition (Long COVID after test-verified SARS-CoV-2 infection) vs broad control definition (population control): https://my.locuszoom.org/gwas/192226/?token=09a18cf9138243db9cdf79ff6930fdf8

Broad case definition (Long COVID after any SARS-CoV-2 infection) vs broad control definition: https://my.locuszoom.org/gwas/826733/?token=c7274597af504bf3811de6d742921bc8

Strict case definition vs strict control definition (individuals that had SARS-CoV-2 but did not develop Long COVID): https://my.locuszoom.org/gwas/793752/?token=0dc986619af14b6e8a564c580d3220b4

Broad case definition vs strict control definition: https://my.locuszoom.org/gwas/91854/?token=723e672edf13478e817ca44b56c0c068

## Supporting information

Authorship info

## Data Availability

We have made the results of these GWAS meta-analyses publicly available for variants passing post-meta-analysis filtering for minor allele frequency >=1% and effective sample size >1/3 of the maximum effective sample size for each meta-analysis. These can be accessed at LocusZoom, where the associations can be visually explored and the summary statistics exported for further scientific discovery.
1) Strict case definition (Long COVID after test-verified SARS-CoV-2 infection) vs broad control definition (population control)
2) Broad case definition (Long COVID after any SARS-CoV-2 infection) vs broad control definition
3) Strict case definition vs strict control definition (individuals that had SARS-CoV-2 but did not develop Long COVID)
4) Broad case definition vs strict control definition

https://my.locuszoom.org/gwas/192226/

https://my.locuszoom.org/gwas/793752/

https://my.locuszoom.org/gwas/826733/

https://my.locuszoom.org/gwas/91854/

