## Supplementary Figures and Table Legends for "Genome-wide Association Study of Long COVID"

### Supplementary Information

Extended Figures **2**
Fig. S1. Frequency of Long COVID symptoms. 2
Fig. S2. Manhattan plots of each of the four Long COVID GWAS meta-analyses. 3
Fig. S3. Chromosome 6 lead variant across the contributing studies and ancestries in GWAS meta-analyses. 5
Fig. S4. Minor allele frequency of lead variant across ancestries. 6
Fig. S5. Principal component (PC) projection. 7
Fig. S6. Expression quantitative trait loci (eQTL) across tissues. 8
Fig. S7. Colocalization analyses of Long COVID with *FOXP4* eQTL, lung cancer, and COVID-19 hospitalization. 9
Fig. S8. Phenome-wide association study (PheWAS) of the lead variant. 11
Extended Tables **12**
Table S1. Sample size for studies contributing to the Long COVID HGI Data Freeze 4. 12
Table S2. Subjects and methods used in each contributing cohort. 12
Table S3. Genome-wide significant variants in GWAS meta-analysis of Long COVID. 12
Table S4. The list of significant variants spanning the genomic region chr6:41,512,355-41,537,458. 12
Table S5. Long COVID *FOXP4* haplotypes in European ancestry. 13
Table S6. Colocalization results for Long COVID with Lung Cancer (Biobank Japan), COVID hospitalization risk (COVID-19 HGI) and 49 GTEx v8 tissue eQTLs at *FOXP4* locus. 13
Table S7. Phenome-wide association study (PheWAS) of Long COVID lead variant in Biobank Japan. 13
Table S8. Roadmap epigenomic marks at lead variants. 14
Table S9. Transcription factor binding sites at lead variants. 14
Table S10. Traits investigated in the genetic correlation and Mendelian randomization analyses. 14
Table S11. Genetic correlations between potential risk factors, biomarkers, and diseases with Long COVID. 14
Table S12. Mendelian randomization of potential risk factors, biomarkers, and diseases with Long COVID. 14
Table S13. Mendelian randomization estimating causal association from SARS-CoV-2 infection and COVID-19 hospitalization on Long COVID. 14
Table S14. Harmonized association statistics for MR exposures and outcomes. 14
References 15

### Extended Figures

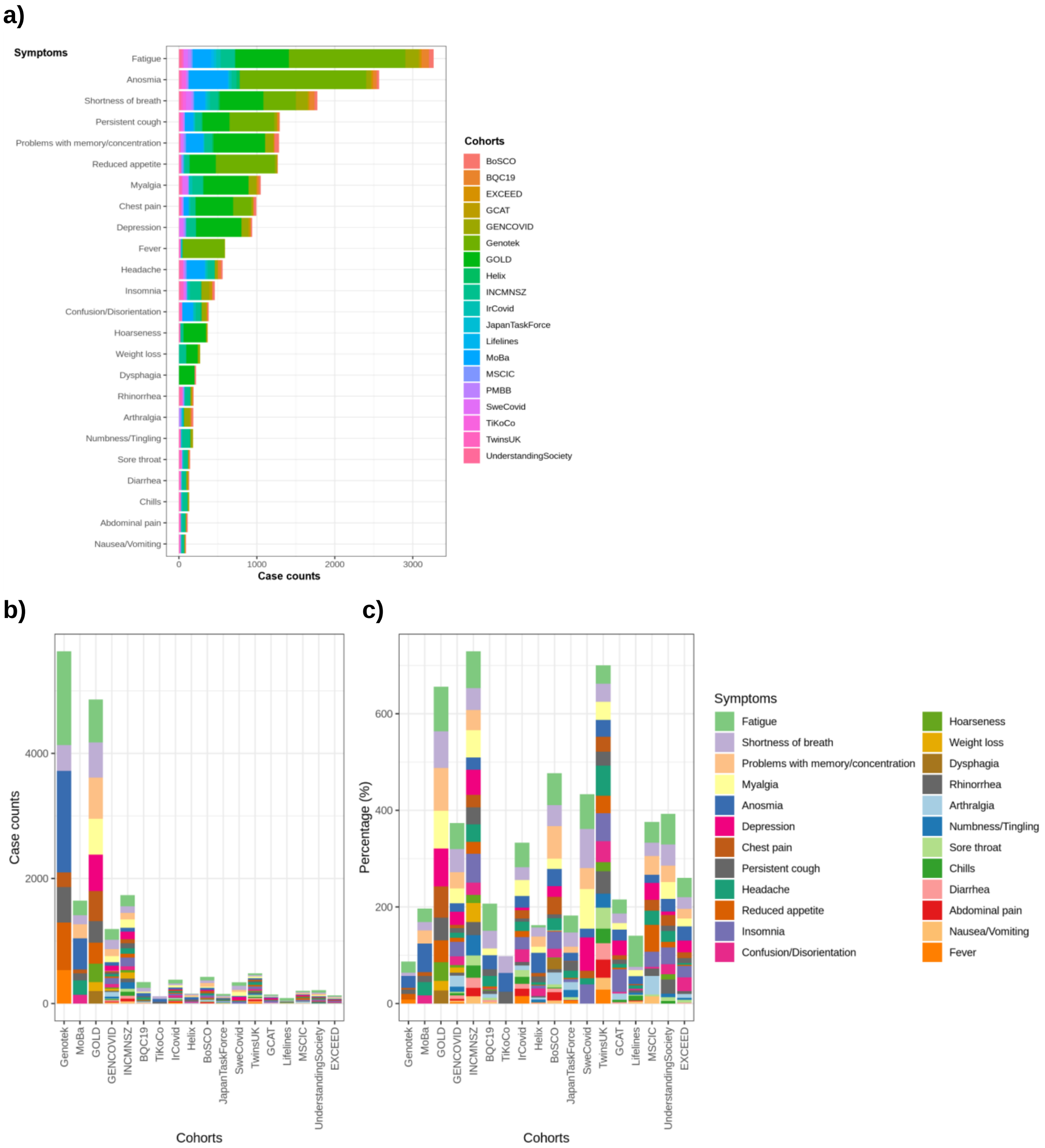


**Fig. S1. Frequency of Long COVID symptoms.**

**a)** Total case counts per each symptom, with contributing cohorts separated by colours.
**b)** Case counts and **c)** percentages of cases with each symptom, stratified by cohorts.
(Note that an individual may have several symptoms, thus the percentages do not total into 100%.)

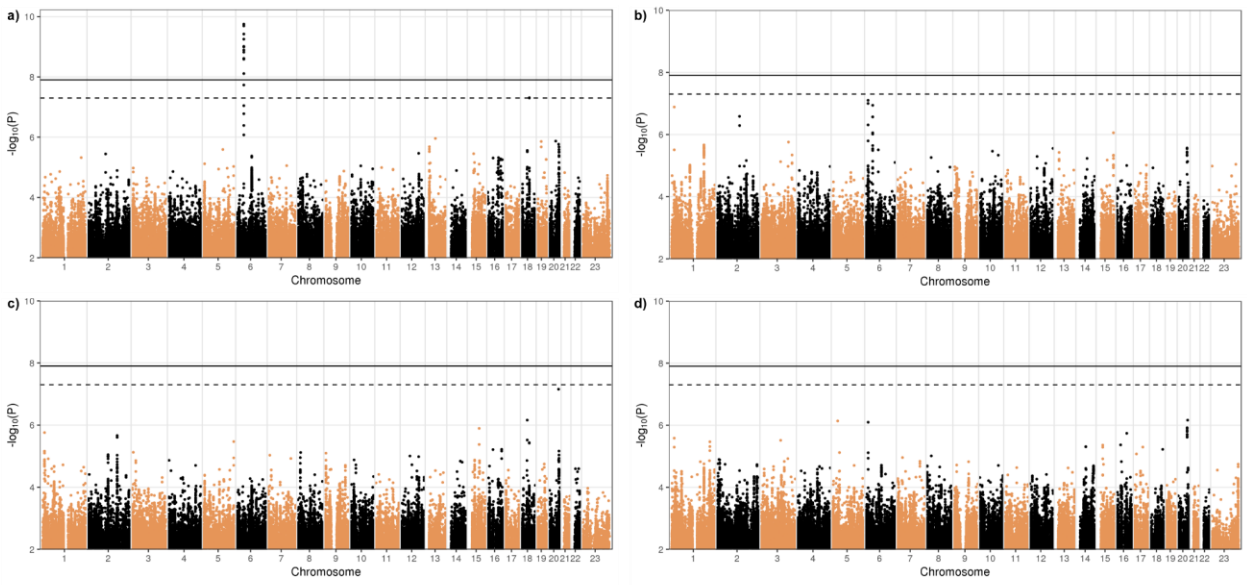


**Fig. S2. Manhattan plots of each of the four Long COVID GWAS meta-analyses.**

Manhattan plots of **a)** Long COVID after test-verified SARS-CoV-2 infection (strict case definition, N = 3,018) compared to all other individuals in each data set (population controls, broad control definition, N = 994,582), **b)** Long COVID after any (test-verified, physician-diagnosed, or self-report) SARS-CoV-2 infection (broad case definition, N = 6,450) compared to population controls (broad control definition, N = 1,093,995), **c)** Long COVID after test-verified SARS-CoV-2 infection (strict case definition, N = 3,018) compared to those recovered within three months after test-verified SARS-CoV-2 infection (strict control definition, N = 37,935), and **d)** Long COVID after any (test-verified, doctor-diagnosed or self-report) SARS-CoV-2 infection (broad case definition, N = 6,450) compared to those recovered within three months after any SARS-CoV-2 infection (strict control definition, N= 46,208). A genome-wide significant association with Long COVID (strict case and broad control definition) was found in the chromosome 6, upstream of the *FOXP4* gene (chr6:41515652:G:C, GRCh38, rs9367106, as the lead variant; $P = 1.76\times10^{-10}$, Bonferroni $P = 7.06\times10^{-10}$, increased risk with the C allele, OR = 1.63, 95% CI: 1.40-1.89). Horizontal lines indicate genome-wide significant thresholds before ($P < 5\times10^{-8}$, dashed line) and after ($P < 1.25\times10^{-8}$, solid line) Bonferroni correction over the four Long COVID meta-analyses.

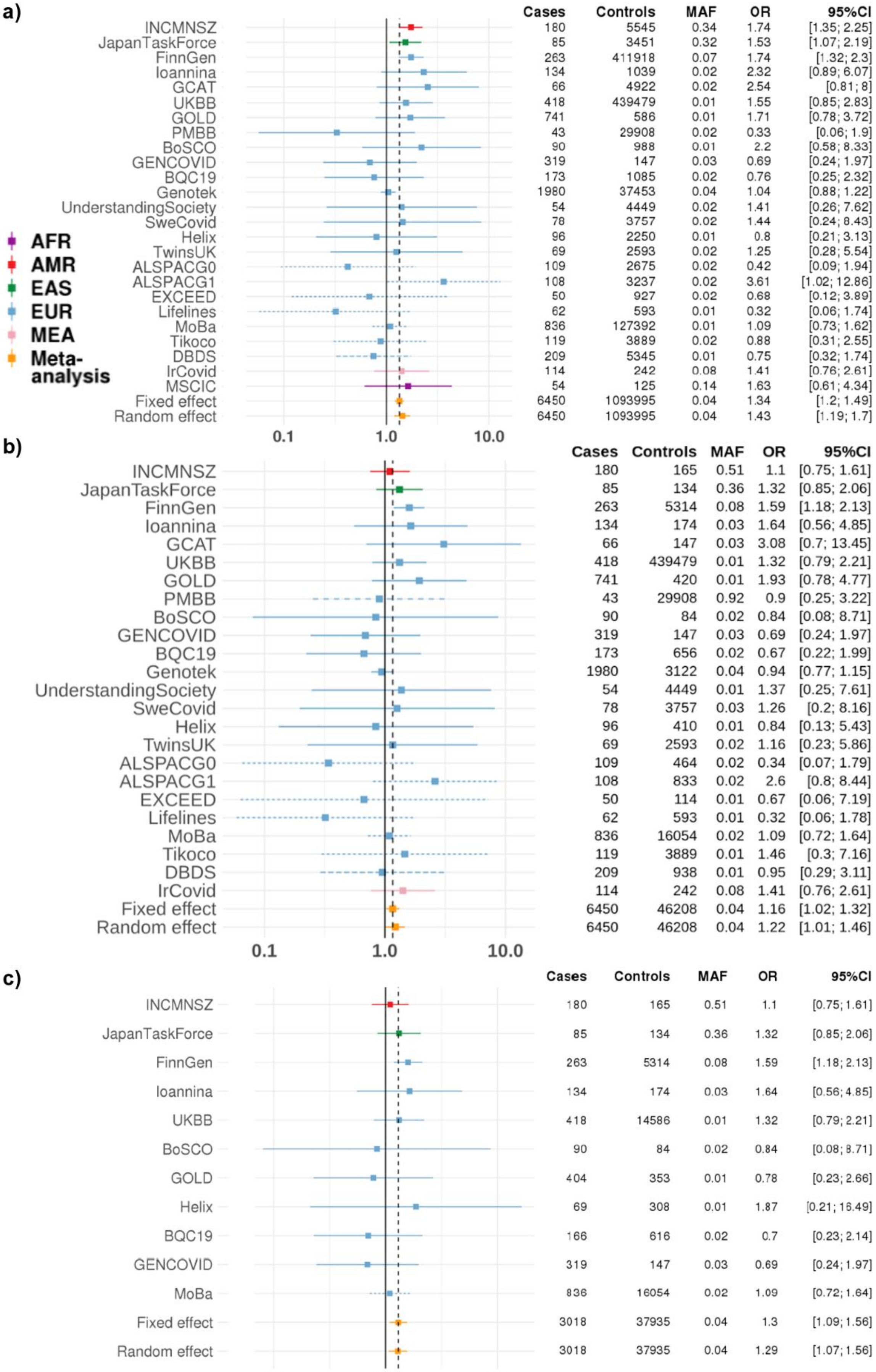
a)
Cases Controls MAF OR 95%CI
INCMNSZ 180 5545 0.34 1.74 [1.35; 2.25]
JapanTaskForce 85 3451 0.32 1.53 [1.07; 2.19]
FinnGen 263 411918 0.07 1.74 [1.32; 2.3]
Ioannina 134 1039 0.02 2.32 [0.89; 6.07]
GCAT 66 4922 0.02 2.54 [0.81; 8]
UKBB 418 439479 0.01 1.55 [0.85; 2.83]
GOLD 741 586 0.01 1.71 [0.78; 3.72]
PMBB 43 29908 0.02 0.33 [0.06; 1.9]
BoSCO 90 988 0.01 2.2 [0.58; 8.33]
GENCOVID 319 147 0.03 0.69 [0.24; 1.97]
BQC19 173 1085 0.02 0.76 [0.25; 2.32]
Genotek 1980 37453 0.04 1.04 [0.88; 1.22]
UnderstandingSociety 54 4449 0.02 1.41 [0.26; 7.62]
SweCovid 78 3757 0.02 1.44 [0.24; 8.43]
Helix 96 2250 0.01 0.8 [0.21; 3.13]
TwinsUK 69 2593 0.02 1.25 [0.28; 5.54]
ALSPACG0 109 2675 0.02 0.42 [0.09; 1.94]
ALSPACG1 108 3237 0.02 3.61 [1.02; 12.86]
EXCEED 50 927 0.02 0.68 [0.12; 3.89]
Lifelines 62 593 0.01 0.32 [0.06; 1.74]
MoBa 836 127392 0.01 1.09 [0.73; 1.62]
Tikoco 119 3889 0.02 0.88 [0.31; 2.55]
DBDS 209 5345 0.01 0.75 [0.32; 1.74]
IrCovid 114 242 0.08 1.41 [0.76; 2.61]
MSCIC 54 125 0.14 1.63 [0.61; 4.34]
Fixed effect 6450 1093995 0.04 1.34 [1.2; 1.49]
Random effect 6450 1093995 0.04 1.43 [1.19; 1.7]
AFR
AMR
EAS
EUR
MEA
Meta-analysis
0.1
1.0
10.0
b)
Cases Controls MAF OR 95%CI
INCMNSZ 180 165 0.51 1.1 [0.75; 1.61]
JapanTaskForce 85 134 0.36 1.32 [0.85; 2.06]
FinnGen 263 5314 0.08 1.59 [1.18; 2.13]
Ioannina 134 174 0.03 1.64 [0.56; 4.85]
GCAT 66 147 0.03 3.08 [0.7; 13.45]
UKBB 418 439479 0.01 1.32 [0.79; 2.21]
GOLD 741 420 0.01 1.93 [0.78; 4.77]
PMBB 43 29908 0.92 0.9 [0.25; 3.22]
BoSCO 90 84 0.02 0.84 [0.08; 8.71]
GENCOVID 319 147 0.03 0.69 [0.24; 1.97]
BQC19 173 656 0.02 0.67 [0.22; 1.99]
Genotek 1980 3122 0.04 0.94 [0.77; 1.15]
UnderstandingSociety 54 4449 0.01 1.37 [0.25; 7.61]
SweCovid 78 3757 0.03 1.26 [0.2; 8.16]
Helix 96 410 0.01 0.84 [0.13; 5.43]
TwinsUK 69 2593 0.02 1.16 [0.23; 5.86]
ALSPACG0 109 464 0.02 0.34 [0.07; 1.79]
ALSPACG1 108 833 0.02 2.6 [0.8; 8.44]
EXCEED 50 114 0.01 0.67 [0.06; 7.19]
Lifelines 62 593 0.01 0.32 [0.06; 1.78]
MoBa 836 16054 0.02 1.09 [0.72; 1.64]
Tikoco 119 3889 0.01 1.46 [0.3; 7.16]
DBDS 209 938 0.01 0.95 [0.29; 3.11]
IrCovid 114 242 0.08 1.41 [0.76; 2.61]
Fixed effect 6450 46208 0.04 1.16 [1.02; 1.32]
Random effect 6450 46208 0.04 1.22 [1.01; 1.46]
0.1
1.0
10.0
c)
Cases Controls MAF OR 95%CI
INCMNSZ 180 165 0.51 1.1 [0.75; 1.61]
JapanTaskForce 85 134 0.36 1.32 [0.85; 2.06]
FinnGen 263 5314 0.08 1.59 [1.18; 2.13]
Ioannina 134 174 0.03 1.64 [0.56; 4.85]
UKBB 418 14586 0.01 1.32 [0.79; 2.21]
BoSCO 90 84 0.02 0.84 [0.08; 8.71]
GOLD 404 353 0.01 0.78 [0.23; 2.66]
Helix 69 308 0.01 1.87 [0.21; 16.49]
BQC19 166 616 0.02 0.7 [0.23; 2.14]
GENCOVID 319 147 0.03 0.69 [0.24; 1.97]
MoBa 836 16054 0.02 1.09 [0.72; 1.64]
Fixed effect 3018 37935 0.04 1.3 [1.09; 1.56]
Random effect 3018 37935 0.04 1.29 [1.07; 1.56]
0.1
1.0
10.0

**Fig. S3. Chromosome 6 lead variant across the contributing studies and ancestries in GWAS meta-analyses.**

Long COVID lead variant from the meta-analysis with strict case and broad control definition (see **Fig. 2**), rs9367106 (solid line). If missing, imputed by the variant with the highest linkage disequilibrium (LD) with the lead variant for illustrative purpose. Dotted line: rs12660421 (r = 0.98 in European in 1000G+HGDP samples[1]), Long-dashed line: rs1886814 (r = 0.65, for one cohort [DBDS]), Dashed line: rs1886817 (r = 0.52 for one cohort [PMBB] in **c**)). For the imputed variants, beta was weighted by multiplying by the LD correlation coefficient (r = 0.98 or 0.65). Ancestries marked by colours: AFR, African; AMR, Admixed American; EAS, East Asian; EUR, European; MEA, Middle Eastern.

**a)** Long COVID with broad case and broad control definition.

**b)** Long COVID with broad case and strict control definition.

**c)** Long COVID with strict case and strict control definition.

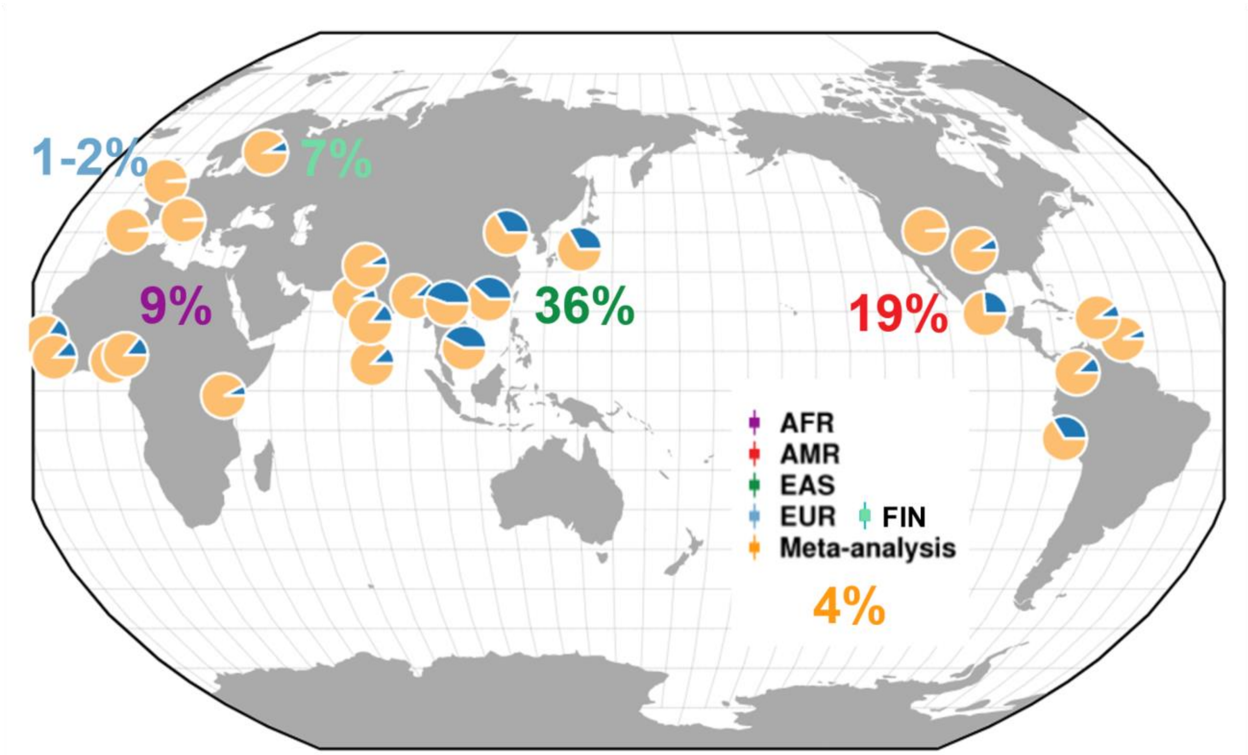


**Fig. S4. Minor allele frequency of lead variant across ancestries.**

Frequency of the Long COVID risk allele (rs9367106-C, marked with blue in pie charts; G allele in yellow) across different ancestries (Geography of Genetic Variants Browser, v0.4 beta, https://popgen.uchicago.edu/ggv/?data=%221000genomes%22&chr=6&pos=41483390)[2]. Long COVID risk allele frequency in populations included in our Long COVID meta-analyses (AFR = African, AMR = Admixed American, EAS = East Asian, EUR = European) marked with ancestry-coloured numbers (gnomAD v3.1.2, https://gnomad.broadinstitute.org/variant/6-41515652-G-C?dataset=gnomad_r3)[2,3]

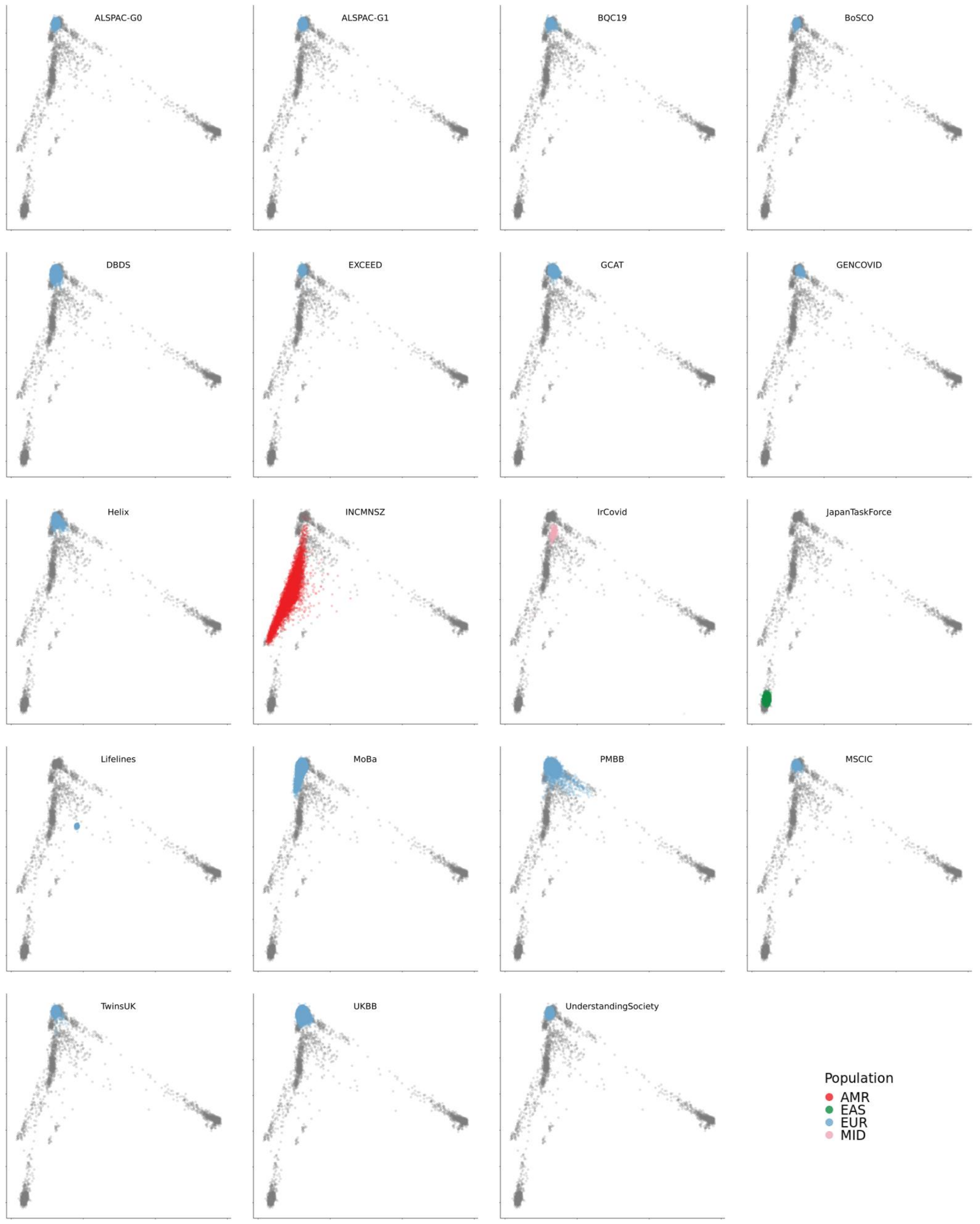


**Fig. S5. Principal component (PC) projection.**

Projection of 1000 Genomes genetic principal components 1 (x-axis) and 2 (y-axis) into studies contributing to the meta-analyses. Each study’s samples are colored based on ancestry (population), with 1000 Genomes samples of all ancestries colored in grey.

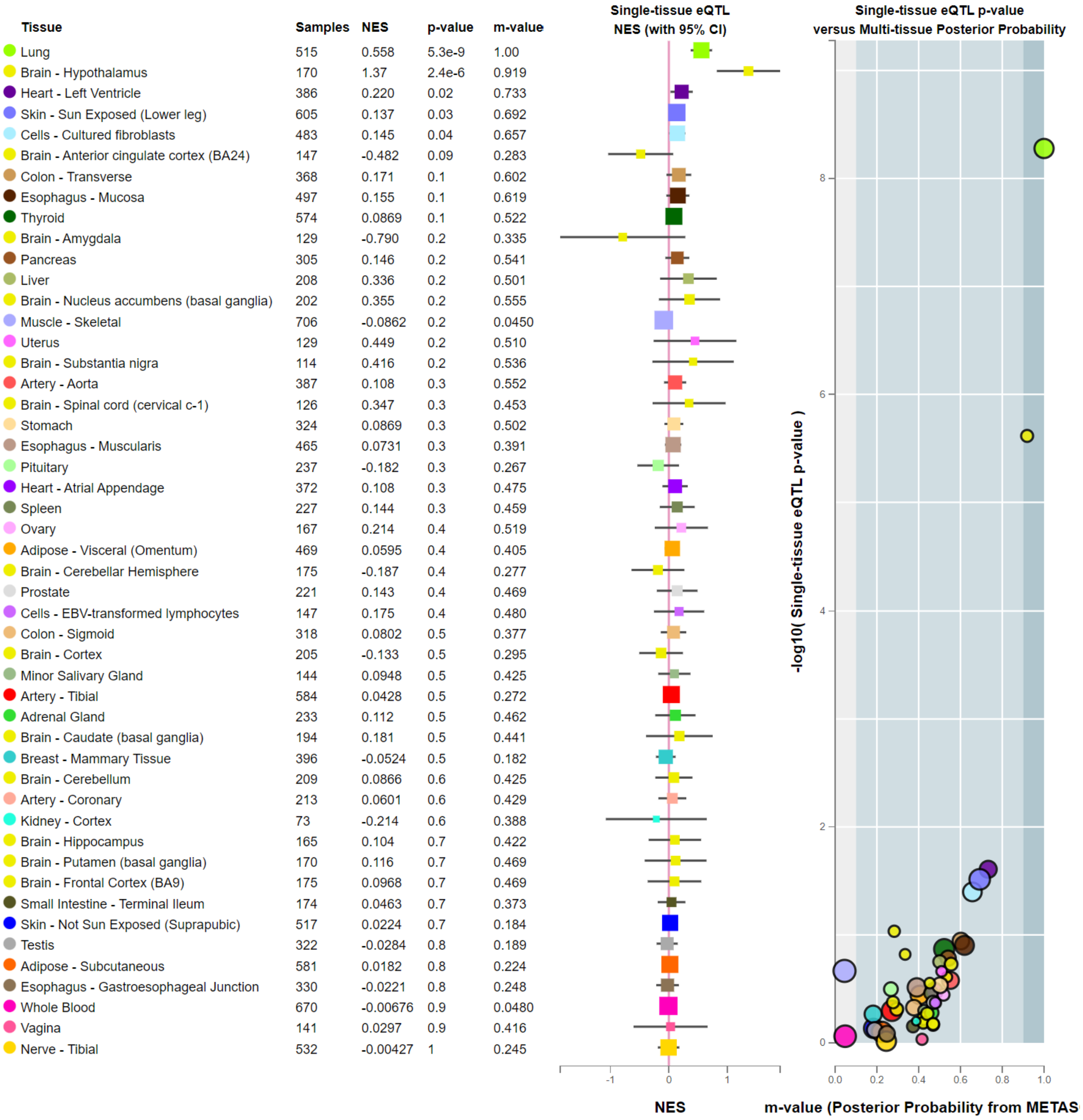

| Tissue | Samples | NES | p-value | m-value |
|---|---|---|---|---|
| Lung | 515 | 0.558 | 5.3e-9 | 1.00 |
| Brain - Hypothalamus | 170 | 1.37 | 2.4e-6 | 0.919 |
| Heart - Left Ventricle | 386 | 0.220 | 0.02 | 0.733 |
| Skin - Sun Exposed (Lower leg) | 605 | 0.137 | 0.03 | 0.692 |
| Cells - Cultured fibroblasts | 483 | 0.145 | 0.04 | 0.657 |
| Brain - Anterior cingulate cortex (BA24) | 147 | -0.482 | 0.09 | 0.283 |
| Colon - Transverse | 368 | 0.171 | 0.1 | 0.602 |
| Esophagus - Mucosa | 497 | 0.155 | 0.1 | 0.619 |
| Thyroid | 574 | 0.0869 | 0.1 | 0.522 |
| Brain - Amygdala | 129 | -0.790 | 0.2 | 0.335 |
| Pancreas | 305 | 0.146 | 0.2 | 0.541 |
| Liver | 208 | 0.336 | 0.2 | 0.501 |
| Brain - Nucleus accumbens (basal ganglia) | 202 | 0.355 | 0.2 | 0.555 |
| Muscle - Skeletal | 706 | -0.0862 | 0.2 | 0.0450 |
| Uterus | 129 | 0.449 | 0.2 | 0.510 |
| Brain - Substantia nigra | 114 | 0.416 | 0.2 | 0.536 |
| Artery - Aorta | 387 | 0.108 | 0.3 | 0.552 |
| Brain - Spinal cord (cervical c-1) | 126 | 0.347 | 0.3 | 0.453 |
| Stomach | 324 | 0.0869 | 0.3 | 0.502 |
| Esophagus - Muscularis | 465 | 0.0731 | 0.3 | 0.391 |
| Pituitary | 237 | -0.182 | 0.3 | 0.267 |
| Heart - Atrial Appendage | 372 | 0.108 | 0.3 | 0.475 |
| Spleen | 227 | 0.144 | 0.3 | 0.459 |
| Ovary | 167 | 0.214 | 0.4 | 0.519 |
| Adipose - Visceral (Omentum) | 469 | 0.0595 | 0.4 | 0.405 |
| Brain - Cerebellar Hemisphere | 175 | -0.187 | 0.4 | 0.277 |
| Prostate | 221 | 0.143 | 0.4 | 0.469 |
| Cells - EBV-transformed lymphocytes | 147 | 0.175 | 0.4 | 0.480 |
| Colon - Sigmoid | 318 | 0.0802 | 0.5 | 0.377 |
| Brain - Cortex | 205 | -0.133 | 0.5 | 0.295 |
| Minor Salivary Gland | 144 | 0.0948 | 0.5 | 0.425 |
| Artery - Tibial | 584 | 0.0428 | 0.5 | 0.272 |
| Adrenal Gland | 233 | 0.112 | 0.5 | 0.462 |
| Brain - Caudate (basal ganglia) | 194 | 0.181 | 0.5 | 0.441 |
| Breast - Mammary Tissue | 396 | -0.0524 | 0.5 | 0.182 |
| Brain - Cerebellum | 209 | 0.0866 | 0.6 | 0.425 |
| Artery - Coronary | 213 | 0.0601 | 0.6 | 0.429 |
| Kidney - Cortex | 73 | -0.214 | 0.6 | 0.388 |
| Brain - Hippocampus | 165 | 0.104 | 0.7 | 0.422 |
| Brain - Putamen (basal ganglia) | 170 | 0.116 | 0.7 | 0.469 |
| Brain - Frontal Cortex (BA9) | 175 | 0.0968 | 0.7 | 0.469 |
| Small Intestine - Terminal Ileum | 174 | 0.0463 | 0.7 | 0.373 |
| Skin - Not Sun Exposed (Suprapubic) | 517 | 0.0224 | 0.7 | 0.184 |
| Testis | 322 | -0.0284 | 0.8 | 0.189 |
| Adipose - Subcutaneous | 581 | 0.0182 | 0.8 | 0.224 |
| Esophagus - Gastroesophageal Junction | 330 | -0.0221 | 0.8 | 0.248 |
| Whole Blood | 670 | -0.00676 | 0.9 | 0.0480 |
| Vagina | 141 | 0.0297 | 0.9 | 0.416 |
| Nerve - Tibial | 532 | -0.00427 | 1 | 0.245 |



#### Fig. S6. Expression quantitative trait loci (eQTL) across tissues.

We show cross-tissue eQTL signals for rs12660421 to allow comparison of signals across tissues (https://gtexportal.org/home/snp/rs12660421). Tissues are sorted by eQTL P value. NES = normalized effect size. m-value = a posterior probability value for each variant-gene pair and tissue tested i.e. the probability that the eQTL effect exists in the given tissue, given the profile of eQTL effects across all investigated tissues (m-value ≥ 0.9 considered as significant)[4].

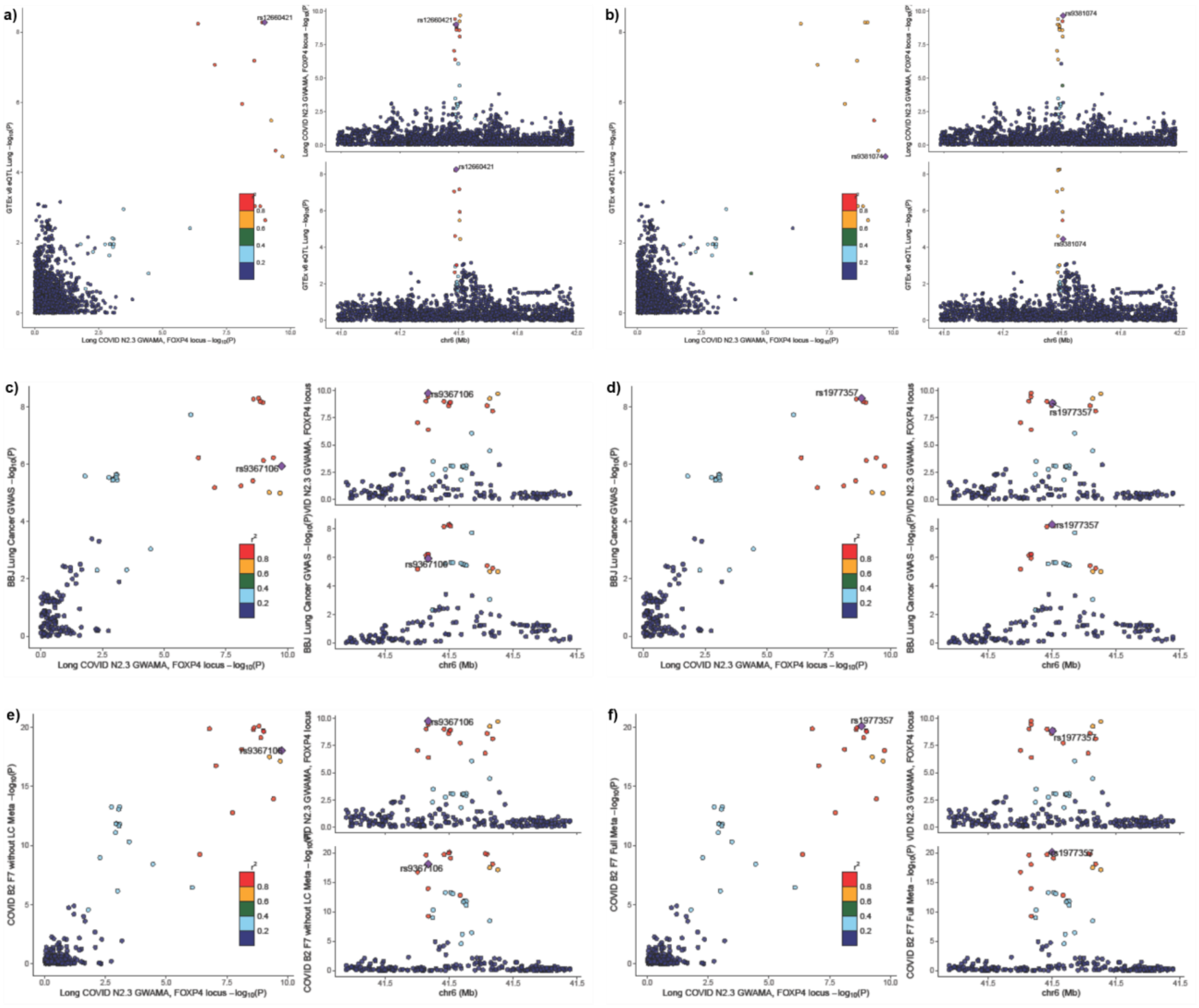


**Fig. S7. Colocalization analyses of Long COVID with *FOXP4* eQTL, lung cancer, and COVID-19 hospitalization.**

(See also **Extended Table S6**)

**a-b)** Long COVID association results with GTEx v8 *FOXP4* expression association results in lung tissue (posterior probability (pp) of shared association = 0.98) in the *FOXP4* locus. Colocalization analysis using eQTL data from GTEx v8 tissue type and Long COVID association data. Plots illustrate -$log_{10}$ P-value for Long COVID (x-axis) and for *FOXP4* expression in the Lung (y-axis), regional association of the FOXP4 locus variants with Long COVID, and regional association of the *FOXP4* variants with RNA expression measured in the lung in GTEx. Variants are coloured by 1000 Genomes European-ancestry LD $r^2$ with **a)** the lead variant (rs12660421) for *FOXP4* expression in lung tissue and **b)** variant (rs9381074) representing the most significant Long COVID variant overlapping the GTEx v8 dataset.

**c-d)** Long COVID association results and Biobank Japan lung cancer association results (pp = 0.91) in the *FOXP4* locus. Plots illustrate -$log_{10}$ P-value for Long COVID (x-axis) and for lung cancer (y-axis), regional association of the *FOXP4* locus variants with Long COVID, and regional association of the *FOXP4* variants with lung cancer. Variants are coloured by 1000 Genomes European-ancestry LD $r^2$ with **c)** the lead variant for Long COVID (rs9367106) and **d)** for lung cancer (rs1977357).

**e-f)** Long COVID association results and COVID-19 hospitalization association results (pp = 0.97) in the *FOXP4* locus. Plots illustrate $-\log_{10}$ P-value for Long COVID (x-axis) and for COVID-19 (y-axis), regional association of the *FOXP4* locus variants with Long COVID, and regional association of the *FOXP4* variants with COVID-19 hospitalization. Variants are coloured by 1000 Genomes European-ancestry LD $r^2$ with **e)** the lead variant (rs9367106) for Long COVID and **f)** for COVID-19 hospitalization (rs1977357).

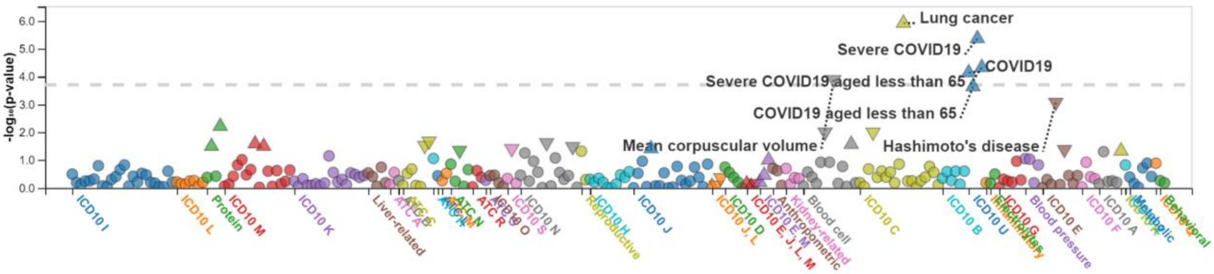


**Fig. S8. Phenome-wide association study (PheWAS) of the lead variant.**

Variant-level PheWAS analysis of the Long COVID lead variant rs9367106 (chr6:41515652:G:C, GRCh38) and all traits (N = 262) in Biobank Japan (https://pheweb.jp/variant/6:41483390-G-C). Significant associations with lung cancer and COVID-19 (P values above the dashed line significant after Bonferroni correction (0.05/262 = $1.9\times10^{-4}$)). (See **Table S7** for all associations with $P < 0.05$.)

### Extended Tables

**Tables in separate file**
LongCOVID_HGI_SupplTables.xlsx

**Table legends**

#### Table S1. Sample size for studies contributing to the Long COVID HGI Data Freeze 4.

Strict case definition = Long COVID after test-verified SARS-CoV2 infection, broad case definition = Long COVID after any SARS-CoV-2 infection. Strict control definition = individuals that had SARS-CoV-2 but did not develop Long COVID, broad control definition = population control i.e. all individuals in each study that did not meet Long COVID criteria.
AFR = African, AMR = Admixed American, EAS = East Asian, EUR = European, MEA = Middle Eastern.
See **Table S2** for more detailed information on the subjects and methods used in each study.

#### Table S2. Subjects and methods used in each contributing cohort.

Information on the subjects (age, sex, hospitalization, Long COVID diagnosis dates) and methods used (phenotype definition, Institutional Review Board (IRB), informed consent, genotype chips, quality control, imputation panel, and ancestry definition) in each study.

#### Table S3. Genome-wide significant variants in GWAS meta-analysis of Long COVID.

**a)** Meta-analysis of 11 studies with strict case definition and broad control definition (see **Table S1** legend or **Methods** for definitions). P values marked with bold were significant after Bonferroni-correction over the four meta-analyses ($P < 5\times10^{-8} / 4 = 1.25 \times 10^{-8}$). Results shown also for leave-most-significant-study-out analysis (LMSO), where the study with the most significant association for each SNP has been excluded and the meta-analysis run with all the other studies for that particular SNP.

#### Table S4. The list of significant variants spanning the genomic region chr6:41,512,355-41,537,458.

The list of 15 variants which span the genomic region chr6:41,512,355-41,537,458, located upstream of the *FOXP4* gene. These variants had a similar effect size to the lead variant, rs9367106, and P values less than $5\times10^{-7}$.

CHR: chromosome, POS: position, EA: effect allele, NEA: non-effect allele, beta/se/pvalue: effect sizes, standard errors, and p-values from the GWAS using the strict case definition (N = 3,018) and the broad control definition (N = 994,582), R_eur/R_sas/R_eas/R_amr/R_afr: LD r value with the lead variant; rs9367106. LD value was calculated using individuals of Europeans in the Human Genome Diversity Project[5] and 1000 Genome Project[6,7]. CADD_phred: PHRED-scaled Combined Annotation Dependent Depletion (CADD) score (a tool for scoring the deleteriousness of single nucleotide variants and insertion/deletion variants in the human genome)[8].

#### Table S5. Long COVID *FOXP4* haplotypes in European ancestry.

Haplotypes constructed using variants in LD ($r^2$>0.5) with the Long COVID lead variant (rs9367106) among Europeans from the 1000 Genomes Project[6]. **a)** Count of individuals and frequency of each haplotype. Main Long COVID risk haplotype in bold. **b)** rsID, chromosomal position (Genome Reference Consortium Human Build 38, GRCh38), and frequency of the alleles for each variant.

#### Table S6. Colocalization results for Long COVID with Lung Cancer (Biobank Japan), COVID hospitalization risk (COVID-19 HGI) and 49 GTEx v8 tissue eQTLs at *FOXP4* locus.

Analysis was performed using the colic R package (v5.1.0.1) in a 1Mb region surrounding the lead variant rs9367106 (chr6:41,015,652-42,015,652). N_SNPs is the number of variants overlapping between Long COVID and the other trait (listed in the Trait column) in the analysis region. PP.HX.abf represent the posterior probability of the approximate Bayes factor for different hypotheses: H0 (no causal variant identified in either dataset), H1 (causal variant identified in trait 1 only - Long COVID), H2 (causal variant identified in trait 2 only - other trait listed in the Trait column), H3 (two distinct causal variants, one for Long COVID and one for the other trait), and H4 (one common causal variant for both Long COVID and the other trait).

#### Table S7. Phenome-wide association study (PheWAS) of Long COVID lead variant in Biobank Japan.

Variant-level PheWAS analysis of the Long COVID lead variant rs9367106 (chr6:41515652:G:C, GRCh38) and all traits (N = 262) in Biobank Japan (https://pheweb.jp/variant/6:41483390-G-C, accessed 18.4.2023) (**Fig. S5**). All associations with $P < 0.05$ (without correction for multiple testing) shown. P values significant after Bonferroni correction ($0.05/262 = 1.9\times10^{-4}$) marked with bold, showing significant associations with lung cancer and COVID-19.

#### Table S8. Roadmap epigenomic marks at lead variants.

Roadmap epigenomic marks retrieved from the Vanno portal http://www.mulinlab.org/vportal/index.html at variants with genome-wide significance or in LD (r2 > 0.5) with the Long COVID lead variant (rs9367106).

#### Table S9. Transcription factor binding sites at lead variants.

Transcription factor binding sites based on Chip sequencing data retrieved from ENCODE, Cistrome and Vanno portal for variants with genome-wide significance or in LD (r2 > 0.5) with the Long COVID lead variant (rs9367106).

#### Table S10. Traits investigated in the genetic correlation and Mendelian randomization analyses.

GWAS summary statistics source, publication, consortium, population, and sample size for each of the 38 traits studied in the genetic correlation and Mendelian randomization analyses.

#### Table S11. Genetic correlations between potential risk factors, biomarkers, and diseases with Long COVID.

Results for LDSC genetic correlations shown in the heatmap **Fig. 5a**.

#### Table S12. Mendelian randomization of potential risk factors, biomarkers, and diseases with Long COVID.

Results for causal associations shown in the heatmap **Fig. 5b**.

#### Table S13. Mendelian randomization estimating causal association from SARS-CoV-2 infection and COVID-19 hospitalization on Long COVID.

COVID-19 Host Genetics Initiative (HGI) GWAS meta-analyses were rerun without cohorts contributing to the Long COVID HGI GWAS meta-analyses to remove sample overlap between the exposures and the outcome. Results from each MR method shown. (See also **Fig. 5c-d**.)

#### Table S14. Harmonized association statistics for MR exposures and outcomes.

Detailed information on the data used for MR (**Fig. 5b, Extended Table S12**).
