## Supplementary Methods for "Genome-wide Association Study of Long COVID"

### Supplementary Methods for
### Genome-wide association of Long COVID

#### Table of contents


Supplementary Methods for Genome-wide association of Long COVID 1

- Long COVID Host Genetics Initiative - Inclusion & Ethics 2
- Phenotype definitions 2
  - Codes to extract cases using registry or electronic health record data 2
  - Strict and broad phenotype definitions 3
- Cohort ancestry and description 4
- Data harmonization 4
- GWAS meta-analyses 4
- Expression quantitative trait loci (eQTL) 4
- Colocalization 5
- Cell-type specific *FOXP4* expression 5
- Enhancers, transcription factor binding sites, and active chromatin regions 5
- Phenome-wide association study (PheWAS) 5
- Mendelian Randomisation 5
- Genetic correlation 7
- References 7

#### Long COVID Host Genetics Initiative - Inclusion & Ethics

The Long COVID Host Genetics Initiative (HGI) is a global and ongoing collaboration project to study genetic factors associated with the risk for developing long-term health problems after SARS-CoV-2 infection. The initiative is open to all studies around the world that have data to run Long COVID genome-wide association study (GWAS) using our phenotypic criteria described below. We encourage studies from all around the world and all ancestries to join this collaborative effort.

The phenotypes and research plan have been designed together by the open global working group of the Long COVID HGI. Each contributing local study has collected the data, run their GWAS, and shared their GWAS summary statistics, which have been meta-analysed together by the initiative. Participants provided informed consent to participate in each respective study, with recruitment and ethics following study-specific protocols approved by their respective Institutional Review Boards and studies performed in accordance with the Declaration of Helsinki (Details are provided in **Extended Table S2**). Contributing researchers from each local study have been acknowledged as co-authors.

#### Phenotype definitions

The current World Health Organization definition includes any symptoms that present after COVID-19 and persist for at least three months[1]. We used clinical diagnosis or self-reported Long COVID in agreement with the World health organisation guidelines following criteria "Post COVID-19 condition occurs in individuals with a history of probable or confirmed SARS CoV-2 infection, usually 3 months from the onset of COVID-19 with symptoms and that last for at least 2 months and cannot be explained by an alternative diagnosis" (https://www.who.int/publications-detail-redirect/WHO-2019-nCoV-Post_COVID-19_condition-Clinical_case_definition-2021.1).

To limit study heterogeneity, we used either self-reported or test-verified COVID-19 infection status to define study participants as Long COVID cases. Self-reported SARS-CoV-2 infection was ascertained if the participant had a documented positive SARS-CoV-2 test result (referred to as "test-verified"), or if they had self-reported suspected COVID-19 (for example, by questionnaire; referred to as "reported") or they had SARS-CoV-2 diagnosis codes in EHR. We aimed to use this broader definition of COVID-19 diagnosis, as many affected individuals may miss a documented COVID-19 diagnosis due to the delay in developing an effective test, limited testing capacity at scale, restrictions on the breadth of public testing, and inadequate record-keeping and linkage, amongst other reasons, especially in the early stages of the pandemic[2–4].

##### Codes to extract cases using registry or electronic health record data

In FinnGen, to include cases based on electronic health record (EHR) data, we used the international classification of diseases version 10 (ICD-10) codes U09.9 (Post COVID-19 condition, unspecified) to assign Long COVID.

In UK Biobank, we used the following codes in the primary care data (the data access was Feb 25, 2022)
TPP local codes: Y2b89, Y2b8a, Y2b87, Y2b88
SNOMED CT: 1325161000000102, 1325031000000108, 1325041000000104, 1325181000000106, 1325021000000106, 1325141000000103, 1325081000000107, 1325061000000103, 1325071000000105, 1325051000000101

We acknowledge that the phenotype definition of Long COVID both by questionnaire and by EHR data is likely to become more precise when more is learned about the disease entity.

#### Strict and broad phenotype definitions

We used the following criteria for assigning case control status for Long COVID aligning with the World Health Organization guidelines (Supplementary Methods). Study participants were defined as Long COVID cases if, at least three months since SARS-CoV-2 infection or COVID-19 onset, they met any of the following criteria:

1) presence of one or more self-reported COVID-19 symptoms that cannot be explained by an alternative diagnosis
2) report of ongoing significant impact on day-to-day
3) any diagnosis codes of Long COVID (e.g. post COVID-19 condition, ICD-10 code U09(.9))

Criteria 1 and 2 were applied only to questionnaire-based cohorts, whereas 3 was used in studies with electronic health records (EHR). Detailed phenotyping criteria and diagnosis codes of each study are provided in **Extended Table S2**.

We used two Long COVID case definitions, a strict definition requiring a test-verified SARS-CoV-2 infection and a broad definition including self-reported or clinician-diagnosed SARS-CoV-2 infection (any Long COVID).

We applied two control definitions. First, we used population controls, i.e. everybody that is not a case. Population controls were genetic-ancestry matched individuals who were not defined as Long COVID cases using the above-mentioned questionnaire or EHR-based definition. In the second analysis, we compared Long COVID cases to individuals who had had SARS-CoV-2 infection but who did not meet the criteria of Long COVID, i.e. had fully recovered within 3 months from the infection.

We used in total four different case-control definitions to generate four GWASs as below;

1) Long COVID cases after test-verified SARS-CoV-2 infection vs population controls (strict case definition vs broad control definition)
2) Long COVID cases within test-verified SARS-CoV-2 infection (strict case definition vs strict control definition)
3) Any Long COVID cases vs population controls (broad case definition vs broad control definition)
4) Long COVID cases within any SARS-CoV-2 infection (broad case definition vs strict control definition)

As all contributing studies did not have data for all of the phenotypes, each meta-analysis comprised those studies that had the phenotypes present and had run that particular GWAS and gone through the quality control. The GWAS results using questionnaire (Q) and EHR (E) based phenotypes were combined in the meta-analysing phase. For studies where all subjects had had a test-verified SARS-CoV-2 infection and thus qualified for the strict case definition, we included the GWASs with strict cases in the meta-analyses with broad case definitions. Similarly, for studies where all control subjects had had a SARS-CoV-2 infection and thus qualified for the strict control definition, we included the GWASs with strict controls in the broad control definition meta-analyses.
Thus, the meta-analysis with broad case and control definitions (marked with ‘3’ in the list above) included data from all of our contributing studies, and the other meta-analyses from subsets of the studies. For this reason, the results of these four Long COVID meta-analyses cannot be directly compared, and the differences between them cannot be interpreted directly as caused by e.g. test-verified vs any SARS-CoV-2, or within-COVID or population-controlled analysis.

#### Cohort ancestry and description

24 studies from 16 countries and 6 ancestries contributed data in the GWAS meta-analyses. In Fig. 1., we display the effective sample size of each analysis which we calculated using the formula $((4 \times N_{case} \times N_{control})/(N_{case} + N_{control}))$. Per-study sample sizes across each phenotype are given in **Extended Table S1**. Study-specific information on participants and methods, including ethics and consent, is provided in **Extended Figure S2**.

Each study projected their cohort onto a multi-ethnic genetic principal component space, with pre-computed PC loadings and reference allele frequencies from unrelated samples from the 1000 Genomes Project and the Human Genome Diversity Project. PCA script internally used the PLINK2 --score function with the variance-standardise option and reference allele frequencies (--read-freq). Consequently, each cohort-specific genotype dosage matrix was mean-centred and variance-standardised with respect to reference allele frequencies, but not cohort-specific allele frequencies. We further normalised the projected PC scores by dividing the values by a square root of the number of variants used for projection to account for a subtle difference due to missing variants.

#### Data harmonization

To allow harmonized data across cohorts, we first compared allele frequencies to Gnomad and aligned alleles to gnomAD 3.0. For any cohorts that were not in genome build 38, we used Picard for liftover. As cohorts provided only summary statistics level information, we examined allele frequency by imputation info scores at cohort level. In addition, we examined association statistics by plotting quantile quantile plots for expected and observed P-values in each cohort. Furthermore, we plotted Manhattan plots for each cohort.

#### GWAS meta-analyses

We computed inverse-variance weighted meta-analysis, which is a method that summarises effect sizes across the multiple studies by computing the mean of the effect sizes weighted by the inverse variance in each individual study. We provide the code to perform the meta-analysis at LongCOVID HGI GitHub (https://github.com/long-covid-hg/META_ANALYSIS/). This meta-analysis pipeline is a modified version of the pipeline used for the main COVID HGI analysis (https://github.com/covid19-hg/META_ANALYSIS). We provide Bonferroni-adjusted threshold that accounts for multiple testing of our 4 phenotypes, albeit it might be overly conservative given that the traits we tested were all Long COVID and were correlated with each other and comprised of partially overlapping individuals. Thus, we also report loci ($P < 5\times10^{-8}$) and report the unadjusted P values for each variant. Furthermore, we investigated the heterogeneity between estimates from contributing studies at variant level using Cochran's Q-test. This is calculated for each variant as the weighted sum of squared differences between the effects sizes and their meta-analysis effect, the weights being the inverse variance of the effect size. Q is distributed as a $\chi 2$ statistic with k (number of studies) minus one degree of freedom.

#### Expression quantitative trait loci (eQTL)

For the single (Bonferroni-corrected) genome-wide significant lead variant, rs9367106, we used the GTEx portal (https://gtexportal.org/) to understand if this variant had any tissue-specific effects on gene expression. As rs9367106 was not available in the GTEx database, we first identified a proxy variant, rs12660421 ($r^2 = 0.90$) using all individuals from the 1000 Genomes Project[5], and then performed a lookup in the portal's GTEx v8 dataset[6].

#### Colocalization

In the coloc R package (v5.1.0.1), we performed all colocalization analyses using the *coloc.abf* function, which calculates approximate Bayes factors, with both p1 (prior probability a SNP is associated with trait 1, Long COVID) and p2 (prior probability a SNP is associated with trait 2, the named trait in the table) set to the default 1e-4 and with p12 (prior probability a SNP is associated with both traits) set to the default 1e-5. For the GTEx colocalization, we used the Ensembl Gene ID "ENSG00000137166" (corresponding to *FOXP4*) to import results from the eQTL catalogue's ftp site (ftp://ftp.ebi.ac.uk/pub/databases/spot/eQTL/imported/GTEx_V8/ge/).

#### Cell-type specific *FOXP4* expression

To assess the relevant cell types for *FOXP4*, we evaluated the transcriptional expression in the lung of healthy controls. We downloaded the single-cell type transcriptomic analyses, where we used all cell types in the lung (GSE13014870). We visualized RNA single cell type tissue cluster data (transcript expression levels summarized per gene and cluster), using log10(protein-transcripts per million (pTPM)) values with "corrplot v 0.92" R package.

#### Enhancers, transcription factor binding sites, and active chromatin regions

We performed functional annotation from ENCODE (https://www.encodeproject.org/), Regulome V2 (https://regulomedb.org/)[7], Cistrome (http://cistrome.org/)[8], and Variant annotation portals (http://www.mulinlab.org/vportal/index.html), examining methylation status, transcriptional activity and transcription factor binding at the variants part of the *FOXP4* haplotype. Furthermore, we identified and visualized methylation and active chromatin regions using the WashU Epigenome Browser[9] (ENCFF778NUQ.bam, ENCFF563OCJ.bam, FOXA1:  ENCFF896BCU.bam, ENCFF631DQI.bam, GATA3: ENCFF999YEG.bam, ENCFF498PGZ, EP300: ENCFF217XRA.bam, ENCFF983ZOH.bam) and validated the DNase and Chip sequencing peaks using Bamtools[10].

#### Phenome-wide association study (PheWAS)

To identify other phenotypes associated with the Long COVID lead variant rs9367106, we used the Biobank Japan PheWeb portal (https://pheweb.jp/)[11] to perform a phenome-wide association analysis, as the minor allele frequency of rs9367106 is highest in East Asia.

#### Mendelian Randomisation

Two-sample Mendelian randomization (MR) was employed to estimate causal associations between 38 cardiometabolic, behavioural, and psychiatric traits and Long COVID using the same approach as previously employed for determining causal associations with COVID-19 susceptibility and severity. Exposures included (**Table S10**): Smoking initiation, Ischemic stroke, High-density lipoproteins, CRP, Diastolic blood pressure, Depression, Insomnia symptoms, Height, Coronary artery disease, Schizophrenia, Lupus, Sleep duration, ADHD, Pulse pressure, Systolic blood pressure, Alzheimer's disease, Risk tolerance, Cigarettes per day, Diabetes, Amyotrophic lateral sclerosis, Rheumatoid arthritis, Multiple sclerosis, Heart failure, Bipolar disorder, Low-density lipoproteins, Total cholesterol, Triglycerides, Chronic kidney disease, BMI, Autism spectrum disorder, Platelet count, Parkinson's disease, Asthma, Red blood cell count, Idiopathic pulmonary fibrosis, White blood cell count, eGFR, and 25 hydroxyvitamin D. We also evaluated the causal association between COVID-19 hospitalization, COVID-19 critical illness, and SARS-CoV-2 infection and Long COVID. These exposures were selected based on their potential as COVID-19 risk factors based on their clinical correlation with disease susceptibility, severity, or mortality. For each exposure, the corresponding publication provides information on how it was measured or diagnosed, the units of measurement used, and the statistical models employed to generate variant associations. Where cross-ancestry discovery GWAS were conducted, EUR-ancestry only GWAS summary statistics were obtained and used in downstream analyses.

MR utilizes genetic variants as proxies for environmental exposures to estimate the causal link between an intermediate exposure and a disease outcome. MR can be compared to a "genetic randomized controlled trial," where risk factors or genotypes are randomly assigned from parents to offspring. This random assignment is not influenced by confounding factors that may affect both risk factors and disease and is unaffected by reverse causation. The genetic variants used in MR function as instrumental variables, provided the following assumptions are satisfied: (1) the genetic variants are known to be associated with the exposure (non-zero effect assumption); (2) the genetic variants are not associated with confounders (independence assumption); and (3) the genetic variants are not directly associated with the outcome (exclusion restriction assumption). In a two-sample MR, analyses are conducted using published genome-wide association summary statistics, with the SNP-exposure and SNP-outcome effects obtained from separate GWAS performed on each trait independently. These separate GWAS are assumed to be conducted in the same underlying population and have no sample overlap.

For each exposure, the respective discovery GWAS was used for both instrument selection and effect size determination. Independent genome-wide significant SNPs ($p < 5e-8$) were chosen as genetic instruments through LD clumping using PLINK ($r2 = 0.001$, 10-Mb clumping window, 1000 Genomes EUR LD reference panel). For genetic instruments not present in the Long COVID GWAS's, PLINK was used to find proxy variants in LD ($r2 > 0.8$). Variants without suitable proxy variants were excluded. The exposure and outcome datasets were then harmonized to ensure that a variant's effect corresponded to the same allele, inferring the positive strand based on allele frequencies for palindromic variants. Causal estimates were calculated using fixed-effect inverse variance weighted (IVW) meta-analysis as the primary analysis and weighted median estimator (WME), weighted mode-based estimator (WMBE), MR-Egger regression, and Mendelian randomization pleiotropy residual sum and outlier (MR-PRESSO) as sensitivity analyses. Although IVW offers the highest statistical power for estimating causal associations, it assumes that all variants are valid instruments and may produce biased estimates if the average pleiotropic effect deviates from zero. Sensitivity analyses provide consistent causal effect estimates even when some instrumental variables are invalid but at the expense of reduced statistical power. The global MR-PRESSO test was employed to assess heterogeneity, and the MR-Egger intercept to evaluate horizontal pleiotropy. Robust causal estimates were defined as those significant at an FDR of 5% and either (1) displayed no evidence of heterogeneity (MR-PRESSO global test $P > 0.05$) or horizontal pleiotropy (Egger intercept $P > 0.05$); or (2) in the presence of heterogeneity or horizontal pleiotropy, the WME-, WMBE-, MR-Egger-, or MR-PRESSO-corrected estimates were significant ($P < 0.05$).

Since no significant causal associations were found between Long COVID and the 38 disease, health, and neuropsychiatric phenotypes, sensitivity analyses like LHC-MR or MRLap, which account for sample overlap, were not carried out. To avoid sample overlap between exposure GWASs (here COVID-19 hospitalization and SARS-CoV-2 reported infection) and outcome GWASs (here Long COVID phenotypes), we performed meta-analyses of COVID-19 hospitalization and SARS-CoV-2 reported infection using data freeze 7 of the COVID-19 HGI by excluding studies that participated in the Long COVID (freeze 4) effort.

The Long COVID phenotype dataset was predominantly composed of individuals of European (EUR) ancestry. However, due to the limited effective sample size available for each phenotype, it was not possible to exclude participants of non-EUR ancestry from the Mendelian randomization (MR) analyses. This decision was made in order to maximize the sample size and statistical power of the analyses, which is crucial for obtaining reliable and accurate results. While the inclusion of participants with non-EUR ancestry may potentially introduce population stratification or confounding effects, the lack of robust causal associations precluded us from investigating this further. Furthermore, as the genetic instruments for the exposures were selected entirely from populations of EUR ancestry, we cannot comment on applicability of our findings to other ancestries. However, the findings should be generalizable across other exposure levels and timings.
All statistical analyses were performed using R v.4.0.3. Mendelian randomization analysis was conducted using the 'TwoSampleMR' v.0.5.5 package.

#### Genetic correlation

We used Linkage disequilibrium score regression (LDSC)[12] to estimate genetic correlations between the Long COVID phenotypes and a set of potential risk factors, biomarkers, and diseases that were earlier studied as part of COVID-19 susceptibility and severity[13]. In addition, we computed genetic correlation analysis with COVID-19 susceptibility and severity. We provide the sources for each GWAS summary statistics for these other traits and the association statistics for all exposure variants as part of the supplementary material (**Extended Tables S10 and S14**, respectively).

Furthermore, we compared the differences between the observed genetic correlations of SARS-CoV-2 infection and COVID-19 severity using a *z*-score method[14].
